# Equity in genome sequencing for rare disease diagnosis: a cross-sectional analysis of data from the UK 100,000 Genomes Project

**DOI:** 10.1101/2024.08.12.24311664

**Authors:** Sam Tallman, Loukas Moutsianas, Thuy Nguyen, Yoonsu Cho, Maxine Mackintosh, Dalia Kasperaviciute, Matthew A Brown, Jamie M. Ellingford, Karoline Kuchenbaecker, Matt J Silver

## Abstract

**Background:** Genome sequencing has improved rare disease diagnosis and is now part of routine clinical care in the National Health Service in England. Automated prioritisation pipelines narrow millions of variants per patient to a small subset for clinical review, a process that relies on allele frequency resources that do not fully represent human genetic diversity. We assessed ancestry-related differences in variant prioritisation and diagnostic outcomes in patients from the UK 100,000 Genomes Project.

**Methods:** We analysed 29,405 rare disease probands with genome sequencing and linked clinical outcomes data. We used multivariable regression to assess ancestry-related differences in prioritised variant counts and positive predictive value (PPV), as well as diagnostic yield and frequencies of recorded variants of uncertain significance (VUS). We also evaluated the use of ancestry-stratified allele frequency filters derived from an independent, diverse UK cohort (n = 31,814).

**Results:** Compared with the European group, the East African group had nearly three times more variants prioritised for clinical review (IRR 2·77, 95% CI 2·33–3·29). Other non-European groups also had significantly higher counts. Diagnostic yield was similar across ancestry groups after adjustment. PPV was lower in East African (OR 0·32, 0·22–0·46), West African (0·47, 0·39–0·57), South Asian (0·65, 0·58–0·73), and Middle Eastern (0·68, 0·54–0·86) groups. VUS frequency was highest in the East African group (OR 1·90, 1·17–3·10) and elevated in West African (1·35, 1·05–1·77) and South Asian (1·33, 1·15–1·59) groups. Applying ancestry-stratified allele-frequency filters removed 1·9% of prioritised variants overall—22·8% in the East African group—without loss of diagnostic sensitivity, including 29·5% of recorded VUS in this group.

**Interpretation:** Differences in PPV and VUS frequencies partly reflect limitations of current allele-frequency resources which use broad population groupings that mask within-group diversity. Increased representation of diverse ancestries in reference databases and better estimation of ancestry-appropriate allele frequencies will help reduce inefficiencies and improve equity in variant prioritisation for rare disease diagnosis.

## Introduction

Over 400 million people worldwide are estimated to be living with a rare disease^1^. While it is thought that more than 80% of rare diseases have a genetic component^1^, most patients do not receive a genetic diagnosis after standard testing^2^. By capturing the majority of the genetic variation in an individual, genome sequencing has improved genetic diagnosis rates^3^. However, distinguishing causal pathogenic variants from the millions of benign variants present in the genome remains a key challenge.

The rare diseases arm of the UK 100 000 Genomes Project (100kGP), is the largest study of sequenced rare disease patients and family members to date^4^. As part of this nation-wide programme, variants identified through genome sequencing and prioritised by Genomics England’s^5^ automated variant prioritisation pipeline have aided in the identification of causal pathogenic variants in thousands of previously undiagnosed participants. The project laid the foundation for the introduction of genome sequencing for patients with rare disease into the National Health Service (NHS) in England as part of routine clinical care^6^.

Pathogenic variants causing rare diseases are generally expected to be rare in all populations^7^. Allele frequency estimates from large-scale reference datasets such as the Genome Aggregation Database (gnomAD)^8^ are therefore used to assist in variant prioritisation by excluding variants too common to plausibly cause the disease. However, a large fraction of global genetic diversity remains to be surveyed and there continues to be an overrepresentation of European ancestries in existing reference datasets^9^. As genomic testing for rare disease diagnosis is incorporated into the NHS and other healthcare systems across the world, it is vital to ensure that advances in genomic medicine translate into accurate and equitable care for all.

We investigated how genetic ancestry affected the performance of Genomics England’s rare disease variant prioritisation pipeline in participants from the 100kGP.

## Methods

### Study design

We analysed genome sequencing and linked health data from participants with rare diseases recruited into the 100kGP. Recruitment to the rare disease programme was coordinated by Genomics England in partnership with 16 NHS Genomic Medicine Centres (GMCs) across the UK between 2015 and 2018, with sequencing completed by December 2018 and results from the automated variant prioritisation pipeline (see “Procedures”) returned to NHS clinical laboratories by July 2019. Study protocols for the 100kGP are available <u>online</u> [https://www.genomicsengland.co.uk/initiatives/100000-genomes-project/documentation].

Ethical approval for the 100kGP was granted by the East of England–Cambridge South Research Ethics Committee (Reference:14/EE/1112; IRAS project ID 166046).

This retrospective analysis was restricted to data included in the version 19 main programme release of the National Genomic Research Library (NGRL)^5^, a research database comprising genome sequencing data and NHS health records including participants in the 100kGP. Analyses were conducted within the secure Genomics England Research Environment under project reference RR786, approved through the Genomics England Research Registry. Patient representatives were not involved in the design or analysis of this study.

### Patients

Patients in the 100kGP rare disease programme were recruited across a broad spectrum of rare conditions (defined as <1 in 2 000 births^10^). NHS medical practitioners determined eligibility based on suspicion of a monogenic or oligogenic disorder without a genetic diagnosis after standard investigations. Where possible, biological parents or other relatives were enrolled to support family-based analyses. Phenotypic data were collected at enrolment using terms selected from curated disease-specific data models, supporting the clinician-led classification of probands and affected relatives into standardised disease groups for downstream analysis (Appendix p11).

Demographic information including year of birth, sex, and self-reported ethnicity was collected alongside clinical data. All participants consented to undergo genome sequencing, receive findings relevant to their condition and permitted access to their de-identified genomic and clinical data for research.

### Procedures

As part of the 100kGP’s primary clinical findings pathway for rare disease, single nucleotide variants (SNVs) and small insertions or deletions (indels) identified through genome sequencing of probands and available family members were processed via Genomics England’s automated variant prioritisation pipeline. Full documentation is available <u>online</u> [https://re-docs.genomicsengland.co.uk/gel_rare_disease.pdf] and a schematic overview is provided in Figure 1.

**Figure 1.**
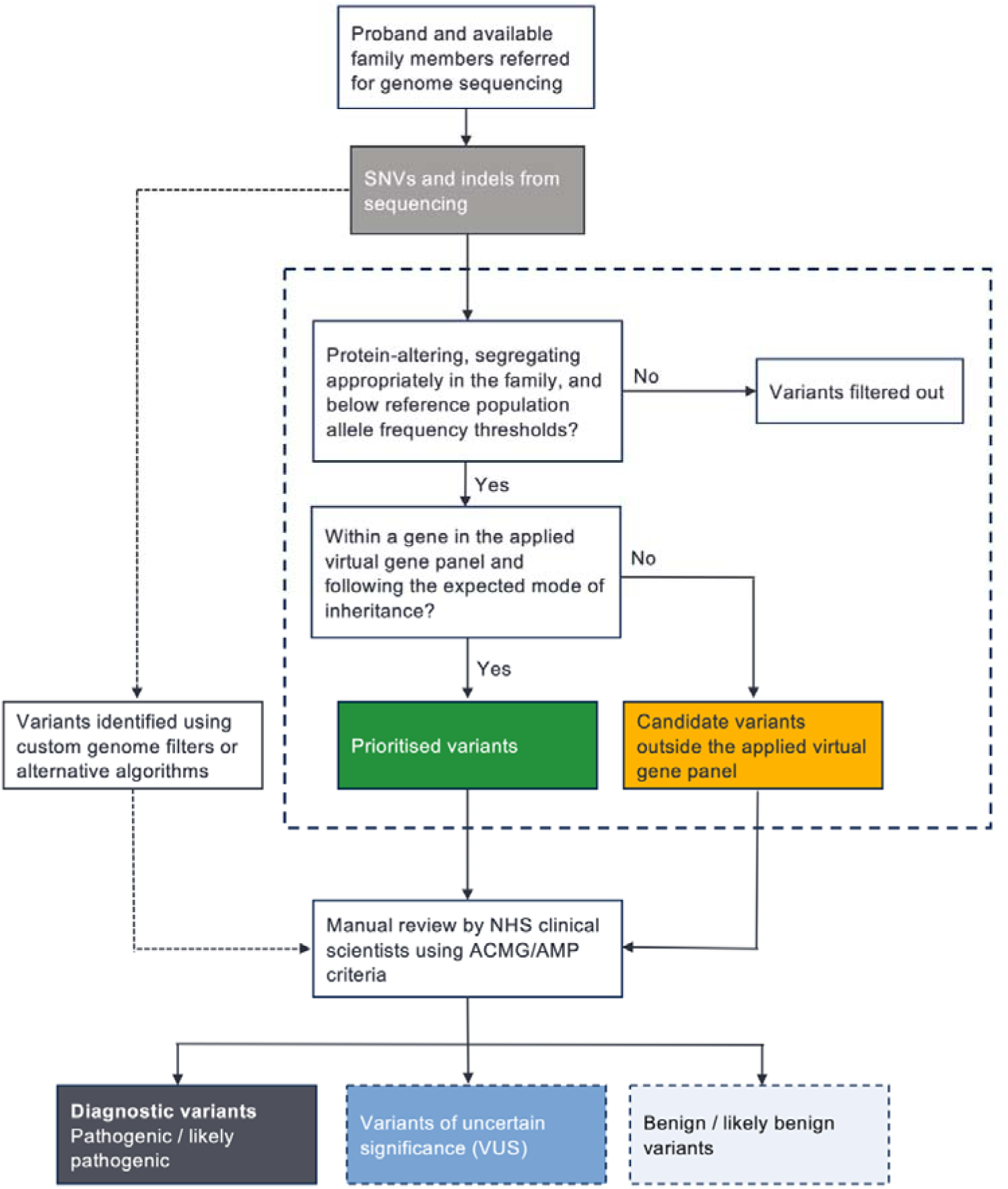
Schematic of the automated variant prioritisation pipeline and manual review process. Single-nucleotide variants (SNVs) and small indels were evaluated by Genomics England’s automated variant prioritisation pipeline (dashed box). Variants were first filtered by predicted protein-altering consequence, familial segregation (where family data were available), and reference population allele frequency thresholds. Further filtering was based on membership of an applied, disease-relevant virtual gene panel and the expected mode of inheritance. *Prioritised variants* passing all the above criteria (green box) were sent for review by NHS clinical scientists. *Candidate variants* outside the applied virtual gene panel (orange box) were also sent but not routinely reviewed. Additional variants could also be identified for review using custom filters or alternative algorithms (dashed arrow). Following interpretation, clinical scientists recorded all *diagnostic variants* that fully or partially explained the proband’s phenotype(s) (black box) in outcomes questionnaires. Non-diagnostic variants, such as *variants of uncertain significance (VUS*, blue box) or benign/likely benign variants (light grey box), could also be recorded but this was not mandatory.

The pipeline aimed to identify potentially causal variants by filtering out variants highly unlikely to cause a rare disease, before prioritising those most likely to explain the proband’s phenotype(s). Filtering retained only predicted protein-altering SNVs and indels that segregated appropriately in the family or that showed strong evidence of being de novo and that occurred at <0·1% frequency (under dominant models) or <1% (under recessive models) in external reference datasets (gnomADv2^8^ and 1000 Genomes^11^) and sequencing technology-matched controls (see Appendix p19 for the applied population frequency filters). Variants passing these criteria were then prioritised if they affected disease-relevant genes and were consistent with the expected mode(s) of inheritance, via the application of expert-assessed virtual gene panels^12^ (continuously updated throughout the programme) selected for the proband’s disease group(s) and clinical presentation. These are referred to as ‘prioritised variants’ hereafter (see Figure 1).

All prioritised variants were reviewed by NHS clinical scientists at the GMCs using American College of Medical Genetics and Genomics and Association of Molecular Pathology (ACMG/AMP) criteria^13^, following best-practice guidelines from the Association for Clinical Genomic Science (ACGS)^14^. Clinical scientists also had the option to extend their review beyond prioritised variants. This included candidate variants that passed the pipeline’s filtering criteria but fell outside the applied virtual gene panels, as well as variants identified through custom filters or alternative algorithms such as Exomiser^15^. For each case, Genomics England collected outcomes from the manual review process through questionnaires submitted by the clinical scientists. These questionnaires recorded whether any of the reviewed variants fully or partially explained the proband’s presenting phenotype(s), and the pathogenic or likely pathogenic variants on which that conclusion was based, hereafter termed ‘diagnostic variants’ (see Figure 1). Reviewed variants not accepted as diagnostic, including those classified as benign, likely benign or of uncertain significance (VUS), could also be recorded but this was not mandatory.

For the present analysis, from the broader cohort of 35 052 probands, we excluded 3543 whose genome sequencing data were aligned to the earlier GRCh37 reference build; 498 without pipeline results or outcome questionnaires available in the NGRL; and 606 whose questionnaires indicated a fully or partially solved case, but with no recorded diagnostic SNVs or indels, such as those based solely on structural variants or chromosomal abnormalities. This left 29 405 probands for downstream analysis.

For this study, we used genome sequencing data to assign each proband to one of eight genetically inferred ancestry groups using a reference dataset curated from UK Biobank^16^, selected for its broad representation of genetic diversity across the UK population. Full methodological details are provided in the Appendix (p4). We use *ancestry group* throughout this Article to refer to these genetic similarity-based groupings, whilst acknowledging that these are simplifications of complex, continuous patterns of genetic ancestry^17^ and that no individuals derive from a single ancestry in any meaningful sense.

Allele frequency estimates in external reference datasets (gnomADv2^8^, 1000 Genomes^11^) used by the variant prioritisation pipeline are stratified by broad continental population groupings (e.g., ‘African’) that do not fully capture African genetic diversity^18^ or that of African diaspora communities in the UK. To explore the potential benefits of using more representative allele frequency estimates, we tested the application of additional allele frequency filters, stratified by the same eight ancestry groups described above (which include distinct East, West, and North African groups; Appendix p9-10). These were derived from a UK cohort of 31 814 genetically unrelated individuals recruited separately for the COVID-19 Genomics Study^19^, that were not enriched for rare diseases. Filtering allele frequencies were adjusted for group-specific sample sizes^20^. We also evaluated additional filters derived from the latest gnomAD release (v4; April 2024) comprising 730 947 exomes and 76 215 genomes, which includes a single ‘African / African American’ reference grouping. Further methodological details are provided in the Appendix (p5-6).

Note, these additional analyses were conducted retrospectively for research purposes only. Results were not returned to NHS clinical teams and did not influence clinical reporting or diagnostic decisions.

### Statistical analysis

We used negative binomial mixed-effects regression to compare the number of prioritised variants across ancestry groups (with all unassigned probands grouped under *remaining participants*), accounting for overdispersion and including random intercepts for the proband’s disease group and handling GMC. Probands with more than one clinically indicated disease group were grouped under *Multiple disorders*. Prespecified covariates, selected based on clinical relevance and prior evidence, included age at recruitment, sex, predicted consanguinity, family structure, and family affection status (Table 1). Probands for whom family affection data was unavailable (2750 probands [9.3%]) were grouped as *unknown* and included in all models.

**Table 1.** Clinical and demographic characteristics for 29 405 analysed probands from the 100kGP rare diseases programme stratified by eight genetically inferred ancestry groups and remaining participants. IQR = interquartile range. *Other family structures include larger family structures or those including non-parental relatives. ^†^ Consanguinity was inferred using the fraction of the genome in long runs-of-homozygosity (ROH)^28^. Categories with less than five individuals (&lt;5) are masked in line with Genomics England reporting practices

|  | European | East Asian | Middle Eastern | Ashkenazi | South Asian | North African | East African | West African | Remaining participants |
| --- | --- | --- | --- | --- | --- | --- | --- | --- | --- |
| **n (% of total)** | 23623 (80.3%) | 124 (0.04%) | 452 (1.5%) | 250 (0.09%) | 3030 (10.3%) | 147 (0.4%) | 159 (0.5%) | 679 (2.3%) | 941 (3.2%) |
| **Age at recruitment** |  |  |  |  |  |  |  |  |  |
| median (IQR) | 26 (8-50) | 30 (10-43) | 26 (8-47) | 51 (27-64) | 14 (6-35) | 17 (7-40) | 20 (9-40) | 33 (12-50) | 13 (6-35) |
| **Reported sex** |  |  |  |  |  |  |  |  |  |
| Female, n (%) | 11542 (48.9%) | 63 (50.8%) | 181 (40.0%) | 130 (52.0%) | 1319 (43.5%) | 71 (48.3%) | 64 (40.3%) | 344 (50.7%) | 419 (44.5%) |
| Male, n (%) | 12081 (51.1%) | 61 (49.2%) | 271 (60.0%) | 120 (48.0%) | 1711 (56.5%) | 76 (51.7%) | 95 (59.7%) | 335 (49.3%) | 522 (55.5%) |
| **Family structure** |  |  |  |  |  |  |  |  |  |
| Singleton, n (%) | 9303 (39.4%) | 69 (55.6%) | 199 (44.0%) | 159 (63.6%) | 939 (31.0%) | 54 (36.7%) | 70 (44.0%) | 373 (54.9%) | 288 (30.6%) |
| Family duo, n (%) | 3363 (14.2%) | 9 (7.3%) | 45 (10.0%) | 12 (4.8%) | 399 (13.2%) | 23 (15.6%) | 30 (18.9%) | 133 (19.6%) | 199 (21.1%) |
| Family trio, n (%) | 8018 (33.9%) | 33 (26.6%) | 143 (31.6%) | 47 (18.8%) | 1189 (39.2%) | 49 (33.3%) | 39 (24.5%) | 121 (17.8%) | 355 (37.7%) |
| Other*, n (%) | 2939 (12.4%) | 13 (10.5%) | 65 (14.4%) | 32 (12.8%) | 503 (16.6%) | 21 (14.3%) | 20 (12.6%) | 52 (7.7%) | 99 (10.5%) |
| **Family disease affection** |  |  |  |  |  |  |  |  |  |
| Relative affected, n (%) | 6365 (26.9%) | 33 (26.6%) | 134 (29.6%) | 81 (32.4%) | 828 (27.3%) | 44 (29.9%) | 43 (27.0%) | 166 (24.4%) | 180 (21.5%) |
| Only proband affected, n (%) | 14901 (63.1%) | 75 (60.5%) | 286 (63.3%) | 122 (48.8%) | 2038 (67.3%) | 93 (63.3%) | 103 (64.8%) | 454 (66.9%) | 606 (72.3%) |
| Unknown, n (%) | 2357 (10.0%) | 16 (12.9%) | 32 (7.1%) | 47 (18.8%) | 164 (5.4%) | 10 (6.8%) | 13 (8.2%) | 59 (8.7%) | 52 (6.2%) |
| **Consanguinity<sup>†</sup>** |  |  |  |  |  |  |  |  |  |
| Consanguineous, n (%) | 136 (0.6%) | <5 | 156 (34.5%) | <5 | 1176 (38.8%) | 46 (31.3%) | 29 (18.2%) | <5 | 21 (2.5%) |
| Possibly consanguineous, n (%) | 45 (0.2%) | <5 | 41 (9.1%) | <5 | 200 (6.6%) | 14 (9.5%) | 11 (6.9%) | <5 | 10 (1.2%) |
| Nonconsanguineous, n (%) | 23442 (99.2%) | 122 (98.4%) | 255 (56.4%) | 247 (98.8%) | 1654 (54.6%) | 87 (59.2%) | 119 (74.8%) | 676 (99.6%) | 807 (96.3%) |

We used logistic mixed-effects regression to compare ancestry group differences in four measures derived from the clinical scientist outcomes questionnaires. Models included random intercepts for disease group and the handling GMC and were adjusted for the same covariates listed above (Table 1). The measures were: (1) diagnostic yield (fully or partially solved case recorded vs none); (2) positive predictive value (PPV; the probability a prioritised variant was recorded as diagnostic); (3) sensitivity (the probability that a diagnostic variant was prioritised; the denominator was all diagnostic variants, including those identified only through extended review beyond the prioritised variants); and (4) VUS frequency (≥1 variant recorded as VUS vs none, per proband).

Likelihood ratio tests (LRTs) assessed the overall significance of ancestry group by comparing multivariable models with ancestry group as a fixed effect against baseline models excluding this term. Adjusted incidence rate ratios (IRRs; for negative binomial models) and adjusted odds ratios (ORs; for logistic models) are reported with 95% confidence intervals (CIs), using the European ancestry group as reference.

Sensitivity analyses compared the PPV and sensitivity of prioritised variants using virtual gene panels applied at the time of sequencing to those using updated versions from January 2024. Stratified analyses examined ancestry group differences across variant consequence categories and inheritance models, as annotated by the variant prioritisation pipeline. Details of these analyses are in the Appendix (p7, p8).

All tests were two-sided, with p < 0·05 denoting statistical significance. Regression analyses were performed in R (version 4.1.1) using glmmTMB^21^ (version 1.1.2.3). Analysis code is available <u>online</u> [https://gitlab.com/genomicsengland/Data_Diversity_Public/rare-diseases-ancestry-paper].

## Results

We analysed 29 405 probands from the 100kGP rare disease programme (Methods). Disorders spanned 218 diseases across 20 broad disease groups. The most common group was neurodevelopmental disorders with 12 003 participants (40·8%) (Appendix p11). Self-reported ethnic group representation broadly matched census data for England (Office of National Statistics 2021) given the cohort’s age profile (Appendix p3, p27).

Participants were assigned to one of eight genetically inferred ancestry groups to capture broad differences in genetic diversity within the cohort (Methods; Appendix p9-10, p28). Those not confidently assigned to any of these eight groups are labelled ‘remaining participants’. Ancestry group assignments and clinical and demographic characteristics included as covariates in downstream multivariable regression models (Methods) are summarised in Table 1.

Across the cohort, a total of 56 642 SNV and indels were prioritised by the automated variant prioritisation pipeline (Methods) and returned to NHS clinical scientists for manual review (see Figure 1). A median of one prioritised variant was returned per proband (interquartile range [IQR] 0–2) with none returned for 17 790 probands (60·5%) (Appendix p12).

The number of prioritised variants returned across ancestry groups ranged from a median of 0 (IQR 0–1) in the Ashkenazi group to 4 (IQR 1–8) in the East African group (Figure 2A). In multivariable negative-binomial regression, ancestry group remained an independent predictor of prioritised variant counts (LRT χ² = 492; p < 0·0001). Compared to the European ancestry group, the adjusted IRR for prioritised variants for the Ashkenazi group was 0·75 (95% CI 0·63–0·89; p = 0·0010), 2·77 (95% CI 2·33–3·29; p < 0.0001) for the East African group, and 1·29–1·79 (all p < 0·0015) for all other ancestry groups (Appendix p14). These differences persisted in an analysis of 5·3 million candidate variants returned outside of the disease-relevant virtual gene panels (LRT χ² = 2 684; p < 0·0001; Figure 1 and Figure 2B) and were not explained by differences in the number of SNV and indels evaluated by the pipeline (Figure 2C). In stratified analyses by variant consequence types and inheritance patterns, missense variants prioritised under inherited dominant models emerged as the principal driver of ancestry group differences (Appendix p29).

**Figure 2.**
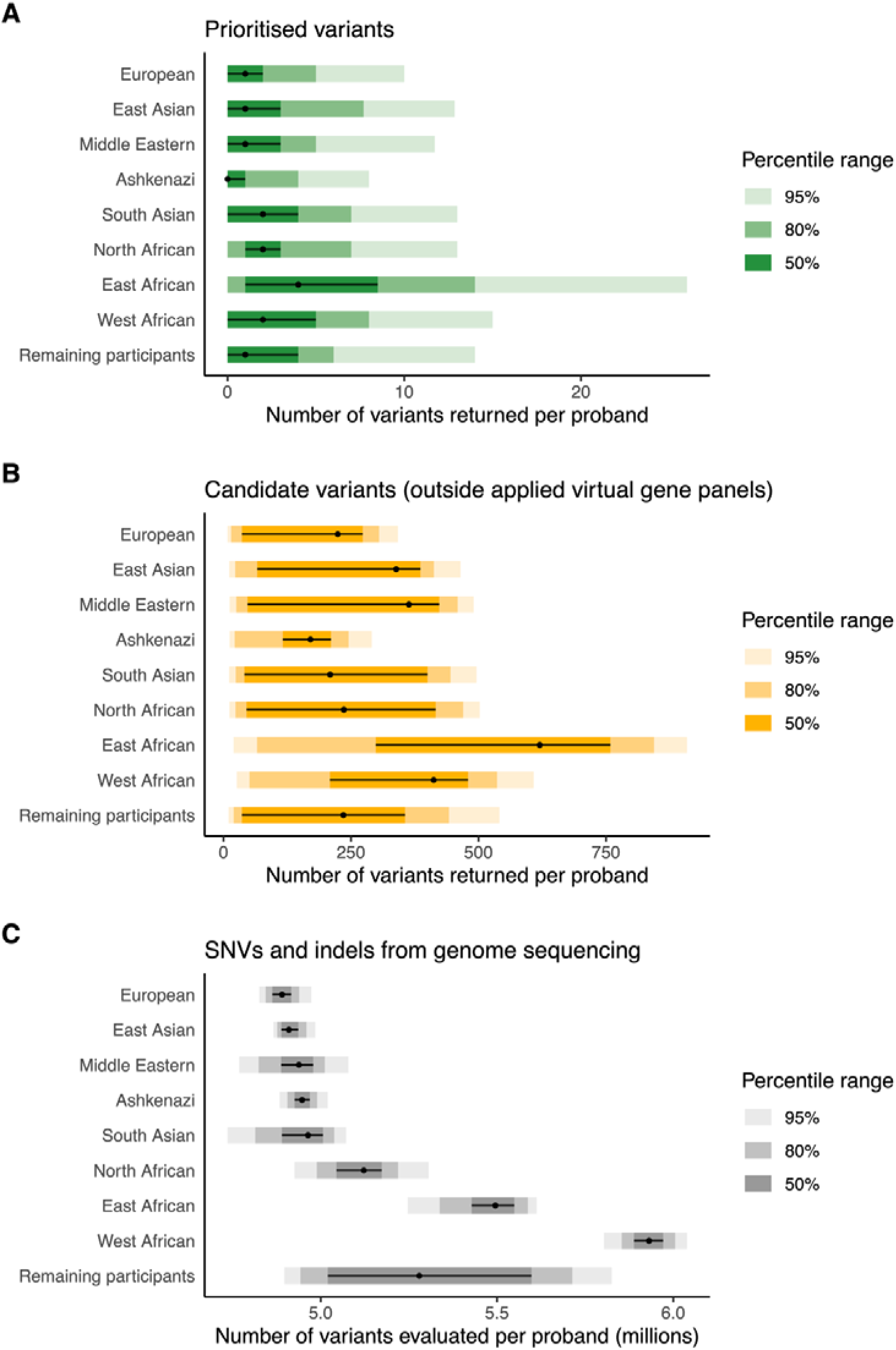
**Distribution of variant counts per proband stratified by genetically inferred ancestry group**. (**A**) Prioritised variants (green). (**B**) Candidate variants returned by the automated pipeline but appearing outside the applied virtual gene panels (orange). (**C**) Total numbers of SNVs and indels from genome sequencing evaluated by the pipeline (grey). Black dots indicate the median per–proband count and black horizontal lines the interquartile range. Shaded bars represent 50th, 80th and 95^th^ percentile ranges. Summary statistics are reported in the Appendix (p12).

Of the probands analysed, 5 662 (19·3%) received a full or partial genetic diagnosis based on SNVs or indels, regardless of whether the variants were prioritised by the automated pipeline or identified through extended review (Methods; Figure 1). Diagnostic yield varied from 18·0% (122/679 probands) in the West African ancestry group to 29·0% (36/124) in the East Asian ancestry group, with 18·3% (4335/23 623) in the European ancestry group (full results in Appendix p13). In multivariable logistic regression models, ancestry group was not independently associated with diagnostic yield (LRT χ² = 11·7; p = 0·1650). In this adjusted model, higher diagnostic yield was associated with younger age, male sex, consanguinity, trio family structure and having an affected relative. Full effect estimates are given in the Appendix (p15).

Of 6506 diagnostic SNVs and indels recorded by clinical scientists, 5262 were among the 56 642 prioritised variants identified across the cohort, yielding an overall sensitivity of 80·9% (5262/6506) and a positive predictive value (PPV) of 9·3% (5262/56 642). Sensitivity was broadly consistent across ancestry groups (range: 91/119 [76·5%] to 32/37 [86·2%]; LRT χ² = 8·4; p = 0.395) (Figure 3A), but PPV varied markedly, ranging from 3·4% (32/955) in the East African group to 13·0% (42/323) in the Ashkenazi group (Figure 3B left). Ancestry group remained an independent predictor of PPV in multivariable logistic regression models (LRT χ² =189; p < 0·0001). Compared to the European ancestry group, the adjusted OR of a prioritised variant being diagnostic was lower in the East African (0·32; 95% CI 0·22-0·46; p < 0·0001); West African (0·47; 0·39-0.57; p < 0·0001); Middle Eastern (0·68; 0·54-0·86; p < 0·0001); South Asian (0·65; 0·58-0·73; p < 0·0001); and remaining participant (0·59; 0·50-0·70; p < 0·0001) groups and higher in the Ashkenazi group (1·45; 1·02-2·06; p = 0·0382) (Figure 3B right; Appendix p17). Similar findings were observed in a sensitivity analysis using virtual gene panels with updated gene-disease associations (Appendix p31).

**Figure 3.**
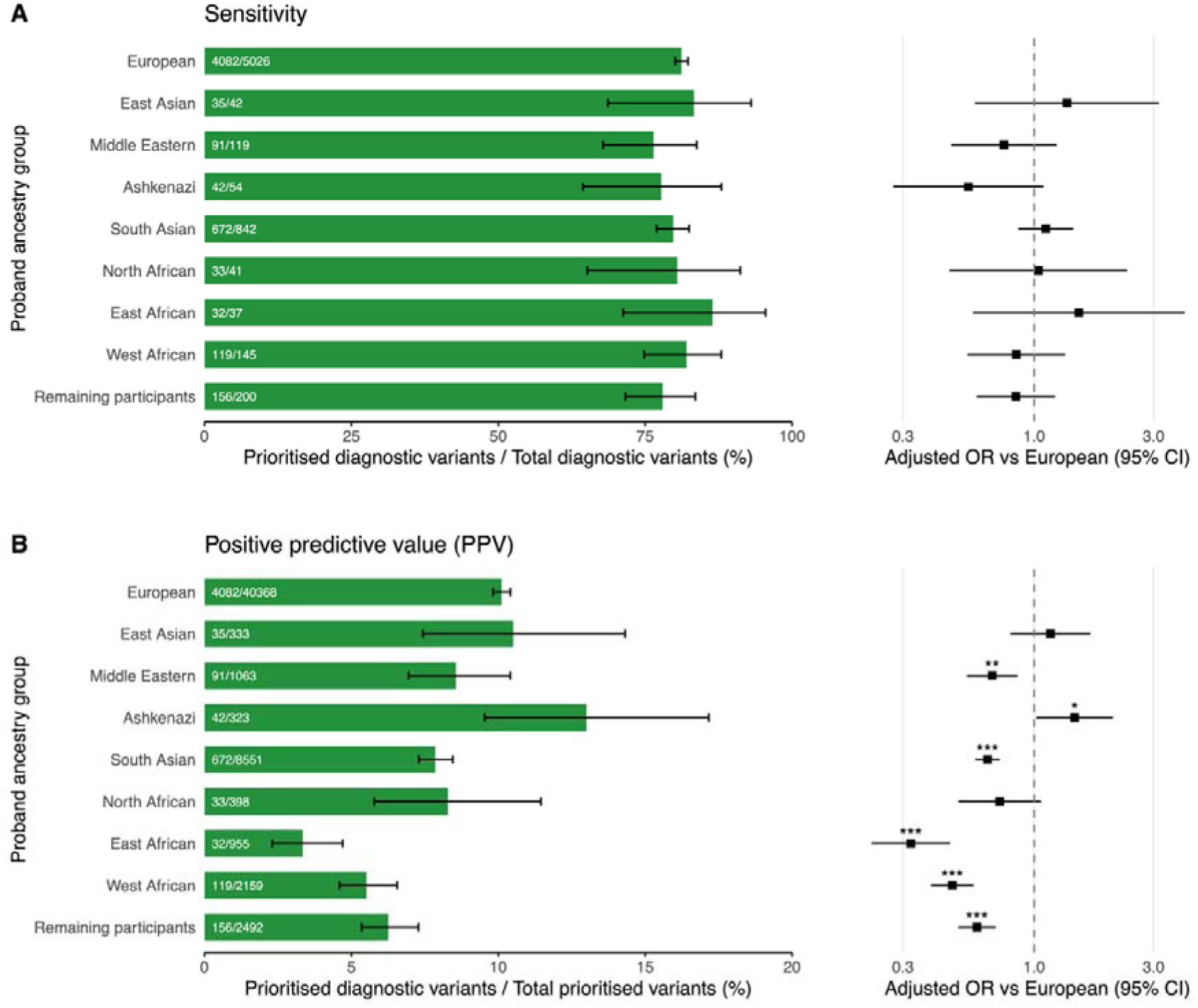
Sensitivity and positive predictive value of variants prioritised by the pipeline stratified by genetically inferred ancestry group. **(A)** Left: proportion of all recorded diagnostic SNVs and indels resulting from prioritised variants (sensitivity) shown as green bars with numerator/denominator labels inside each bar and 95% Clopper–Pearson CIs (whiskers). Right: adjustedLJOR forest plot. **(B)** Left: proportion of prioritised variants recorded as diagnostic (PPV) shown as green bars with numerator/denominator labels inside each bar and 95% Clopper–Pearson CIs (whiskers). Right: adjusted-OR forest plot. Estimates of adjusted OR vs European are from multivariable mixedLJeffects logistic regression models adjusted for age, sex, family structure, familyLJmember affection status, and predicted consanguinity, with random intercepts for disease group and the handling GMC. *** p<0·001; ** p<0·01; * p<0·05). Full regression coefficients, 95% CIs, and p values for all covariates are provided in the Appendix (p15-17).

Overall, 4036 variants were classified as variants of uncertain significance (VUS), with at least one VUS recorded in 9·8% (2889/29 405) of probands. The proportion of probands with at least 1 VUS ranged from 3·2% (8/250) in the Ashkenazi ancestry group to 13·8% (22/159) in the East African ancestry group (Figure 4 left). In multivariable logistic regression models, ancestry group remained an independent predictor of VUS frequency (LRT χ² = 38·8; p < 0·0001). Compared with the European ancestry group, the adjusted OR of having at least one recorded VUS was higher in the East African (OR 1.90; 95% CI 1·17–3·10; p = 0·0097); West African (1·36; 1·05–1·77; p = 0·0217); South Asian (1·35; 1·15–1·59; p = 0·0003) and remaining participant (1·45; 1·17–1·78; p = 0·0005) groups (Figure 4 right, Appendix p18).

**Figure 4.**
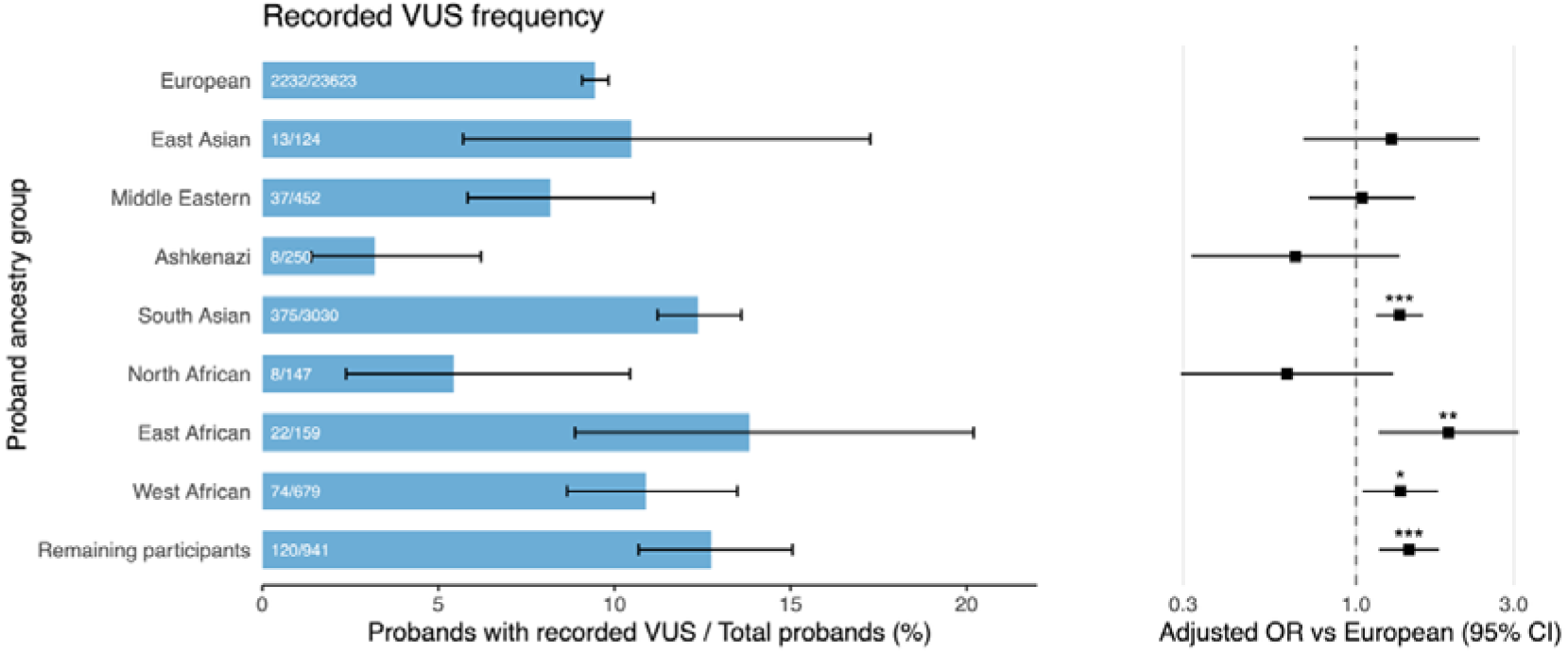
Probands with at least one recorded variant of unknown significance (VUS), stratified by genetically inferred ancestry group. Left: percentage of probands with at least one VUS (light blue bars), with numerator/denominator labels and 95% Clopper–Pearson confidence intervals (CIs; whiskers). Right: Forest plot with adjusted odds ratios (ORs) and 95% CIs from multivariable mixed-effects logistic regression of ancestry group on VUS carrier status, compared to the European ancestry group. Models adjusted for age, sex, family structure, family-member affection status, and consanguinity, with random intercepts for disease group and handling GMC (Methods). *** p<0·001; ** p<0·01; * p<0·05). Full regression coefficients, 95% CIs, and p values for all covariates are provided in the Appendix (p18).

We next assessed if ancestry-stratified allele frequency estimates that reflect the genetic diversity of the 100kGP cohort could improve the PPV of prioritised variants and reduce the burden of recorded VUS. To do this, we re-ran the variant prioritisation pipeline using allele frequency filters derived from a relatively small number (31 814) of diverse genomes from the UK COVID-19 Genomics Study^19^, stratified by the same eight ancestry groups considered throughout our analysis (Figure 5 blue bars; Appendix p20). This removed 1·9% of previously prioritised variants (1070/56 642) without loss of sensitivity (5069/5069, i.e. 100% of diagnostic variants retained). The largest effect was in the East African ancestry group (23·3% [218/955] of prioritised variants removed) and the smallest effect was in the European ancestry group (0·8% [318/40 368] removed). Other ancestry groups saw 1·5% (5/333) to 4·0% (16/398) of variants removed (Figure 5).

**Figure 5.**
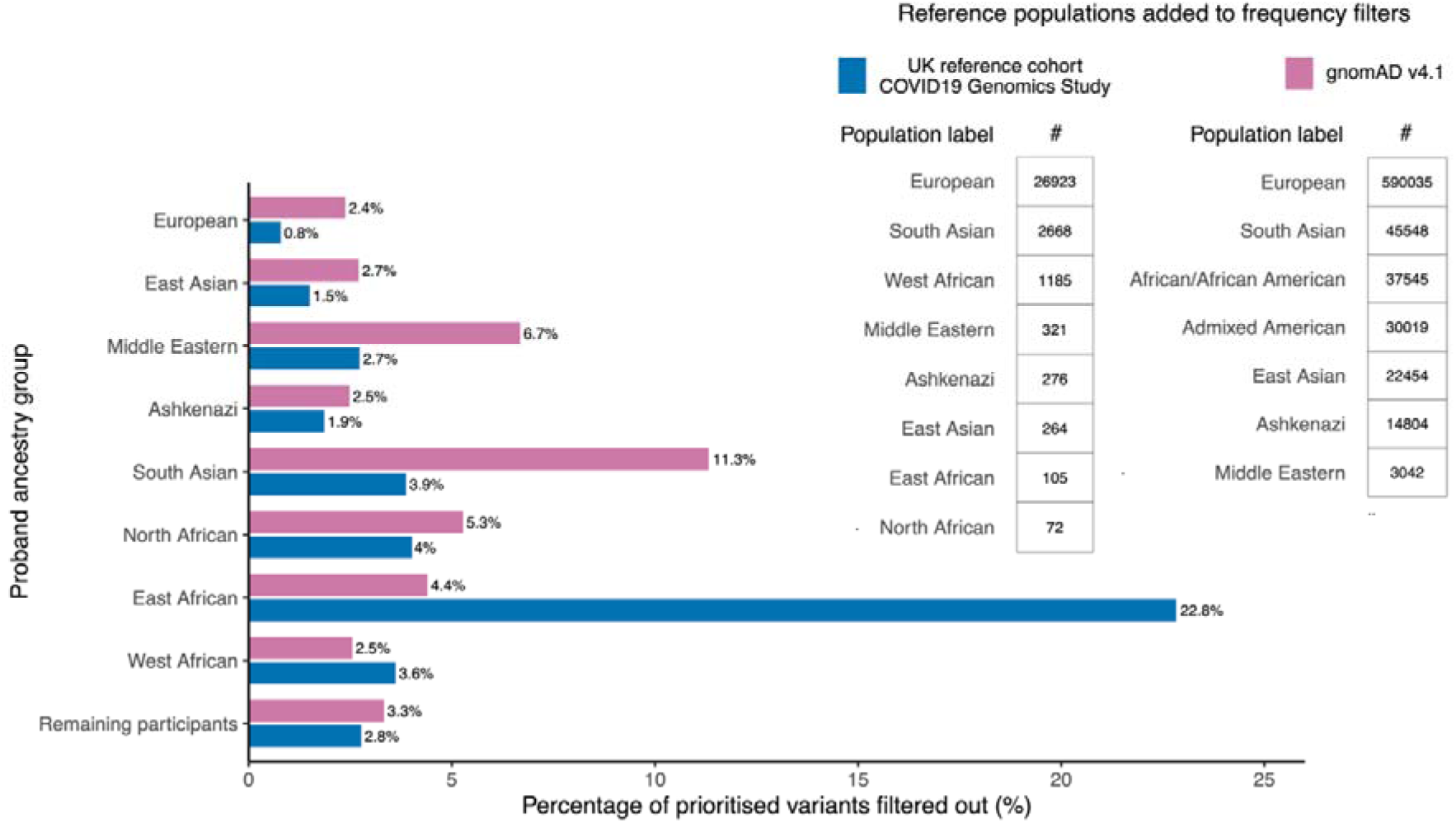
Impact of additional allele frequency filters on prioritised variants across eight genetically inferred ancestry groups. Blue bars show the percentage of prioritised variants removed in each proband ancestry group when additional allele frequency thresholds estimated from the UK COVID-19 Genomics Study (blue) were added to automated pipeline filters (Methods). Magenta bars show the same for allele frequencies derived from gnomADv4.1 (genomes; magenta). Sample sizes for each labelled group used in allele frequency estimation are in the inset tables. Full filter definitions and methodological details, including sample-size adjustments are shown in the Appendix (p5-6).

When substituting these frequency filters with those from gnomADv4^8^, a substantially larger dataset that notably lacks a dedicated East African reference group (Figure 5 magenta bars; Appendix p21), an additional 0·6–7·4% of prioritised variants were removed in all ancestry groups except East and West African, where 18·4% (4·4% [42/955] vs 22·8% [218/955]) and 1·1% (2·5% [55/2159] vs 3·6% [78/2159]) fewer variants were removed respectively. Similar patterns were observed when filtering variants outside of the applied virtual panels (Appendix p34).

33 VUS recorded in the outcome questionnaires were among those filtered out using the ancestry-stratified frequency thresholds from the UK COVID-19 Genomics Study (full list in the Appendix p22-25). 13 of these were identified in probands in the East African ancestry group; 29.5% (13/44) of all recorded VUS in this group. For example, a missense variant (13-114324819-A-G) in *CHAMP1*, initially prioritised under a dominant inheritance model in a proband with intellectual disability and developmental delay, was absent from gnomADv2, and reached a maximum frequency in gnomADv4 of 0·05% (3 of 6084 alleles) in the Middle Eastern group. It was also absent in other groups in the UK COVID-19 Genomics Study cohort but reached a frequency of 2·3% (5 of 210 alleles) in the East African group (Appendix p25). Given the typically de novo and early-onset nature of *CHAMP1*-related disorders , this high frequency likely exceeds the maximum credible allele frequency for pathogenicity, enabling the use of evidence within the current clinical guidelines^14^ to support a benign classification (BS1).

## Discussion

Genome sequencing is increasingly used in the healthcare pathway for patients with rare diseases as a method to inform diagnosis, prognosis and treatment. The accuracy and precision of genomic approaches for rare disease diagnosis depends on the reliable identification and prioritisation of variants underpinning these conditions. In our analysis of data from the 100kGP, the world’s largest cohort of rare disease patients with genome sequencing and linked clinical data, we demonstrate that genetic ancestry affects the performance of an automated variant prioritisation pipeline used to support genetic diagnosis. While diagnostic yield did not differ significantly by ancestry group in multivariable models, we observed significant disparities in the positive predictive value (PPV) of prioritised variants returned for clinical review and in the frequency of recorded variants of uncertain significance (VUS).

These disparities were most pronounced among patients of genetically inferred East African ancestry. For patients in this group, variants prioritised for clinical review were three times less likely to be accepted as diagnostic compared to those in the European ancestry group, and they also had the highest frequency of recorded VUS. Our findings suggest these discrepancies are partly driven by gaps in ancestry-specific allele frequency data in commonly used reference databases such as gnomAD, which aggregate across broad, continental categories that do not adequately capture African genetic diversity^18^. This means that variants common in individuals with East African ancestries are more likely to appear superficially rare, reducing their positive predictive value and inflating the number of VUS. Indeed, when we retrospectively applied new allele frequency filters, including those derived from a small East African ancestry-specific control group (n = 105), we found that 22·8% of prioritised variants in this group could have been excluded from manual review without loss of diagnostic sensitivity. Notably, this included 29·5% of all recorded VUS in this group, highlighting the clinical utility of more representative allele frequency resources and the potential value in periodically re-evaluating VUS as allele frequency resources become more diverse.

Previous studies have reported lower diagnostic yields for rare disease patients with non-European ancestries^23^, whilst other studies have reported no significant ancestry-related differences^24^. Despite observed differences in the number of variants prioritised and returned for clinical review, we found no evidence that genetic ancestry influenced the likelihood of obtaining a genetic diagnosis in our cohort after adjustment for clinical and demographic covariates. This may reflect the robustness of the manual variant review processes. However, we caution against overinterpreting this finding as the variance in diagnostic yield across the diverse spectrum of disease phenotypes represented in the cohort is large^2^, and patient numbers for many conditions are small. Larger, more diverse rare disease cohorts, including the addition of data from patients referred for genome sequencing by the NHS Genomic Medicine Service^6^, may provide the necessary statistical power to identify ancestry-related differences where they exist.

Our study has several limitations. Firstly, the genetically inferred ancestry groups used here do not fully capture the complex, continuous nature of genetic ancestry^17^. Furthermore, diagnostic outcomes may be influenced by unmodelled sociocultural, clinical, and environmental factors^23^ that could be correlated with these ancestry groups. Reported differences, or lack thereof, in diagnostic outcomes should be interpreted in this context. The predominance of patients of European ancestry and the wide range of rare disease phenotypes in the 100kGP also limit the ability to detect possible disease-specific ancestry biases. Moreover, recording of VUS in the outcomes questionnaires was not mandatory and may have varied between centres, so VUS counts may be under-ascertained and should be interpreted with caution. Finally, whilst this study focussed on protein-altering single-nucleotide variants and small indels, emerging evidence highlights the role of non-coding and large structural variants in rare disease pathogenesis^25,26^. Future studies should assess how genetic ancestry influences the prioritisation of all variant types captured by genome sequencing.

In conclusion, the effectiveness of variant prioritisation approaches using genome sequencing data from patients with rare disease remains limited by the lack of diversity in population reference datasets, and by the broad population groupings used to stratify allele frequency estimates. This can lead to an increased number of false positives amongst prioritised variants and a higher burden of VUS. This has consequences for clinical efficiency and patient care, since false positives can increase clinical scientist workload and associated costs, while unresolved VUS can delay decision-making and contribute to uncertainty around treatment and management^27^. Increased representation of diverse genetic ancestries in genomic reference databases and use of ancestry-appropriate allele frequency estimation is essential for achieving

## Data availability

The data supporting the findings of this study are available within the Genomics England Research Environment. To access genomic and clinical data within the Research Environment, researchers must first apply to become a member of either the Genomics England Research Network (academics/healthcare professionals) or the Discovery Forum (industry) via the Genomics England website.

## Funding

The UK Department of Health and Social Care and the EU’s Horizon 2020 Research and Innovation Programme.

## Acknowledgements

This research was made possible through access to data in the National Genomic Research Library, which is managed by Genomics England Limited (a wholly owned company of the Department of Health and Social Care). The National Genomic Research Library holds data provided by patients and collected by the NHS as part of their care and data collected as part of their participation in research. The National Genomic Research Library is funded by the National Institute for Health Research and NHS England. The Wellcome Trust, Cancer Research UK and the Medical Research Council have also funded research infrastructure. We thank the participants in the 100 000 Genomes Project, who made this study possible. We also acknowledge the data contributed by the GenOMICC, REACT and ISARIC4C teams and thank the participants from those studies. The COVID-19 (GenOMICC) study was funded by the Department of Health and Social Care, Illumina, LifeArc, the Medical Research Council (MRC), UKRI, Sepsis Research (the Fiona Elizabeth Agnew Trust), the Intensive Care Society, a Wellcome Trust Senior Research Fellowship (223164/Z/21/Z) a BBSRC Institute Program Support Grant to the Roslin Institute (BBS/E/D/20002172, BBS/E/D/10002070 and BBS/E/D/30002275) and UKRI grants MC_ PC_20004, MC_PC_19025, MC_PC_1905 and MRNO2995X/1. We also acknowledge the National Institute for Healthcare Research Clinical Research Network (NIHR CRN) and the Chief Scientist’s Office (Scotland), who facilitated recruitment into research studies in NHS hospitals, and to the global ISARIC and InFACT consortia. K.K. has received funding from the European Research Council (ERC) under the European Union’s Horizon 2020 research and innovation program (Grant agreement No. 948561).

## Author Contributions

S.T., K.K, and L.M designed the study. S.T. carried out the analysis. L.M., K.K., and M.J.S. supervised the work. S.T., M.J.S., and K.K. wrote the first draft of the manuscript. All authors contributed to discussion of the results and reviewed and edited the manuscript.

## Conflict of Interest Statement

All authors are current or former employees of Genomics England Ltd.

## Role of Funding Source

Funding sources were not involved in the design or analysis of this study.

## Ethnics Committee Approval

Ethical approval for the 100kGP was granted by the East of England–Cambridge South Research Ethics Committee (Reference:14/EE/1112; IRAS project ID 166046). Analyses in this study were conducted within the secure Genomics England Research Environment under project reference RR786, approved through the Genomics England Research Registry.

## Notes

### Competing Interest Statement

All authors are employees of Genomics England Ltd.

### Clinical Protocols

https://www.genomicsengland.co.uk/initiatives/100000-genomes-project/documentation

### Author Declarations

Ethical approval for the 100kGP was granted by the East of England-Cambridge South Research Ethics Committee (Reference:14/EE/1112; IRAS project ID 166046). Analyses in this study were conducted within the secure Genomics England Research Environment under project reference RR786, approved through the Genomics England Research Registry.

### Summary of Updates

The paper has been substantially restructured and re-written to fit a clinical journal style, with updates to the analyses and results using a newer Genomics England data release - as well as updated analyses, Figures, and Tables to improve clinical interpretability. All areas of the paper have thus been substantially re-written to include these updates.

