## Appendix for "Equity in genome sequencing for rare disease diagnosis: a cross-sectional analysis of data from the UK 100,000 Genomes Project"

**Genomics England’s Small Variant Prioritisation Pipeline**

To aid NHS clinical scientist evaluation of primary findings for rare diseases in the 100,000 Genomes Project (100kGP)^1^, single nucleotide variants (SNVs) and indels in families are filtered and prioritised using an automated, virtual panel-based variant prioritisation pipeline, formally referred to as the Rare Disease Small Variant Tiering Process in Genomics England’s documentation. In this study, we analysed **version 5.0 of the pipeline**, specifically as applied to participants in the 100kGP rare disease programme whose genomes were aligned to the **GRCh38 reference build**.

Complete documentation is available online: <https://re-docs.genomicsengland.co.uk/gel_rare_disease.pdf>

Note, in Genomics England’s documentation, returned variants are classified as:

- **Tier 1**: High-confidence de novo or predicted loss-of-function (LoF) variants within genes on the applied virtual gene panels, consistent with the specified inheritance model and passing filtering criteria.
- **Tier 2**: Other protein-altering or splice-region variants (e.g. missense, in-frame indels) in panel genes that pass filtering but do not meet Tier 1 criteria.
- **Tier 3**: Protein-altering variants outside the applied panels, yet also pass segregation pattern and frequency filters

In this study, we did not distinguish between Tier 1 and Tier 2 variants, referring to all variants returned within the applied virtual gene panel(s) by the automated pipeline as prioritised variants (see Figure 1 in main text). Tier 3 variants (i.e. outside panel) were analysed separately. Note, where a variant was returned multiple times for a given proband (e.g. under different applied virtual gene panels or plausible inheritance models), it was counted a single time.

**Table A3** summarises the total number of prioritised and filtered (Tier 3) variants, including the number of unique variants, variant burden per proband, and the proportion of probands with >1 variant, stratified by genetically inferred ancestry group (see Appendix p 3). Variant count distributions per proband across the analysis cohort are visualised in **Figure A1**.

A Table summarising the population frequency filters used by the automated variant prioritisation pipeline (v5; GRCh38) is shown in Table A10.

**Self-reported ethnicity**

Self-reported ethnicity labels were collected for all probands and family members in the UK100kGP at recruitment, using the standard 16+1 categories defined in the NHS data dictionaries (<https://www.datadictionary.nhs.uk>). Among the analysis cohort, these labels were available for 14.6% (4292/29405), with the remainder recorded as 'Not known' or not stated. Additionally, free-text columns for "other" parental ethnicities were available for a subset of participants (2612/29,405; 8.9%).

We assessed the representativeness of self-reported ethnic groups in the cohort relative to the general population in England, by comparing with the 2021 Office for National Statistics (ONS) data (<https://www.ons.gov.uk/peoplepopulationandcommunity/culturalidentity/ethnicity/datasets/populationestimatesbyethnicgroupenglandandwales>).

To harmonise categories, ONS 2021 ethnicity labels were updated to match 16+1 nomenclature (e.g., “Asian, Asian British, or Asian Welsh” → “Asian or Asian British”). ONS categories not directly mapped to the 16+1 set—specifically “White: Gypsy or Irish Traveller,” “White: Roma,” and “Other ethnic group: Arab”—were merged into “White: Any other White background,” “White: Any other White background,” and “Other ethnic groups: Any other ethnic group,” respectively. These mappings were chosen to align with the 16+1 system, but comparisons between ONS data should be considered approximate.

To account for the strong age–ethnicity structure in England, we stratified both datasets by decade (from ages 0 to 70+) and computed the proportion of individuals within each ethnic group per age stratum. Individuals in the UK100kGP cohort with unknown or unstated ethnicity were excluded. Broad agreement was observed across age-stratified trends between the study cohort and ONS data (Figure A2). A notable exception was over-representation of “Asian or Asian British: Pakistani” individuals in deciles under age 30, consistent with elevated consanguinity rates and associated recessive disease burden reported among British Pakistanis. Other modest deviations may reflect demographic or recruitment biases and warrant further investigation beyond the scope of this analysis.

**Genetic ancestry inference**

We assigned all 29 405 rare disease probands in our 100kGP analysis cohort (see Methods; main text) to one of eight ancestry groups using a reference dataset previously curated using data from the UK Biobank^2^ described in Privé et al 2022^3^. This reference dataset was selected due to its broad representation of genetic diversity across the UK population.

The 63,523 sites used for genetic ancestry inference were common, high-quality, linkage disequilibrium (LD) pruned single nucleotide variants (SNVs) previously curated by the Genomics England bioinformatics team. Sites were selected based on the following criteria: (1) biallelic SNVs; (2) minor allele frequency >5% in both the 1000 Genomes Phase III^4^ and the Genomics England multi-sample variant aggregate (aggV2; <https://re-docs.genomicsengland.co.uk/aggv2/>) (3) median genotype quality >30; (4) allele balance ratio >0.9; (5) genotype call rate >90%; (6) exclusion of SNVs within regions of long-range LD; (7) exclusion of ambiguous A/T or G/C SNPs; (8) LD pruning using PLINK^5^ v1.9 with an r^2^ threshold of 0.1 in 500 kb windows; and (9) Hardy-Weinberg Equilibrium p-value >1e^-5^. After applying these filters, 63,523 SNPs were retained. Of these, 55,706 overlapped with the UK Biobank reference dataset from Privé *et al* 2022, which included ~4 million sites, and were used for principal component (PC) projection.

The bigsnpr^6^ R package was first used to project proband genotypes and allele frequencies from an initial set of 21 reference groups across these 55,706 intersecting sites onto the top 16 linkage LD-scaled PCs created from the reference panel. Squared Euclidean distances on the PC space (converted into an approximate F_ST_) were then calculated between each proband and these 21 reference groups, and probands were assigned to any group if F_ST_ < 0.005 (as per the *bigsnpr* manual). To consolidate genetically similar groupings and avoid over-fragmentation, groups were merged and relabelled using a graph-based clustering approach (using a custom R script): an edge was drawn between any two groups if ≥50% of individuals assigned to one were also assigned to the other at F_ST_ < 0.005. Final clusters were defined as connected components in this graph, allowing for transitive merging of overlapping groups. This process yielded 11 merged groups. From this set of merged groups, those with fewer than 100 probands (South America, Japan, Philippines) were then merged and relabelled as ‘remaining participants’ alongside all probands unable to be assigned to any one reference group using the above criteria, likely because of recently mixed genetic ancestries or high divergence from reference groups due to under-representation in the reference dataset.

This resulted in a final set of **eight ancestry groups**, plus those labelled remaining participants. Further details are provided in Table A1, including the reference groups that were merged, the place of birth of individuals in the reference groups, as well as comparisons with 16+1 ethnicity available additional self-reported parental ethnicities where available.

Genetically inferred ancestry groupings aligned with clustering patterns on a genotype PCA previously constructed by the Genomics England bioinformatics team using the GCTA^7^ software applied to the same set of high-quality sites as shown in Figure A3.

**Construction of an ancestry-group stratified allele frequency reference panel and application of filtering allele frequency thresholds**

We constructed an allele frequency reference panel using a multi-sample aggregate of SNVs and small indels from **34,701** whole-genome sequenced participants without rare diseases, curated as part of a UK COVID-19 Genomics study as described in Kousathanas et al. 2021^8^.

To derive a set of genetically unrelated individuals, we applied a kinship coefficient threshold of >0.0442 (3rd-degree relatives), using estimates previously generated by the Genomics England bioinformatics team via the PLINK2 implementation of the KING-robust algorithm^9^. This yielded a set of **33,724** genetically unrelated individuals.

These unrelated individuals were then assigned to one of eight ancestry groups, following the ancestry inference and merging strategy described for the rare disease cohort (see Appendix p4). This resulted in **31,814** participants stratified into one of the eight ancestry groups used for the 100kGP. Final group sizes in the reference panel as shown in Table A11.

To generate ancestry-stratified allele frequency estimates, we used aggregated SNV and indel variant calls from Genomics England's release v5 of the UK COVID-19 Genomics Study, spanning the autosomes and X chromosome. Multi-allelic variants were decomposed into biallelic form. Genotypes were masked if they failed the following thresholds:

- **Autosomal + female X genotypes**: depth (DP) < 10, genotype quality (GQ) < 20, or allele balance (AB) binomial test p < 10⁻³
- **Male X (non-PAR):** DP < 5 or GQ < 20

In total, the aggregated dataset included **505,993,047** SNVs and small indels across **31,814** participants.
For all SNVs and indels, we calculated allele counts (AC) and allele numbers (AN) stratified by ancestry group using PLINK2, separately for the autosomes and X chromosome. In non-pseudo autosomal regions of the X, male genotypes were counted as haploid. Only variants PASS variants with greater than >50% coverage across the dataset were included.

Using this dataset, we calculated conservative, ancestry group-specific estimates of allele frequency using the filtering allele frequency (FAF) approach described by Whiffin et al 2017^10^. We defined FAF as the **lower bound of the 95% confidence interval for the true population allele frequency, given the observed allele count and sample size,** using a Poisson model, with singletons (AC = 1) conservatively assigned a FAF of 0. This provides a cautious estimate of how common a variant could plausibly be, helping to avoid misclassifying variants as too common to be disease-causing when they may only appear at a high frequency due to limited sampling size within a given ancestry group.

We evaluated the potential of this diverse, ancestry-matched reference panel by retrospectively integrating the derived FAF thresholds into the automated variant prioritisation pipeline used in the UK100kGP (Appendix p 2). Specifically, across all 5,719,061 variants (1,974,633 unique; whether within the applied virtual panels or not) that passed the standard filtering criteria during 100kGP—including population frequency thresholds from gnomAD v2, 1000 Genomes, and custom controls (Table A10)—we additionally applied FAF thresholds of 0.001 for variants flagged under recessive inheritance models and 0.0001 for those flagged under dominant models (see Table A11). To maintain consistency with the variant prioritisation pipeline versions currently used by the NHS Genomic Medicine Service (GMS) in the UK (<https://genomicsengland.gitlab.io/rare_disease_analysis/rare-disease-analysis-guide/Mira/>), we retained all variants classified as “Pathogenic” or “Likely Pathogenic” in ClinVar^11^ (as of March 2024), regardless of their observed frequencies. 100kGP cohort-wide FAF distributions of variants originally returned by the pipeline, and the number of such variants that failed these thresholds, are shown in Figures A7 and A8, stratified by reference group and whether the variant was prioritised based on the applied virtual gene panel.

Of note, **33 previously prioritised variants that were recorded as variants of uncertain clinical significance (VUS) in clinical scientist-issued outcomes questionnaires** were filtered out using these ancestry group-specific FAF thresholds. Full details of these variants, including their maximum observed frequencies across ancestry groups, are provided in Table A13.

For comparison, we also derived FAF thresholds from the gnomAD^12^ v4.1 dataset comprising 730 947 exomes and 76 215 genomes into seven populations (Finnish and remaining groups in the gnomAD v4.1 release were excluded from this analysis). Variants failing quality control (QC) filters, showing highly discrepant frequencies between exomes and genomes, or with callable coverage in less than 50% of individuals in the dataset were excluded to minimise the inclusion of technical artefacts and improve the reliability of allele frequency estimates. Using the same Poisson-based approach applied to the UK COVID-19 Genomics Study panel, we calculated population-specific FAFs across SNVs and indels in gnomAD v4.1 spanning the autosomes and X chromosome. These FAF-based thresholds - 0.0001 for dominant and 0.001 for recessive inheritance models (Table A12) - were then applied in addition to the standard filtering criteria used by the automated variant prioritisation pipeline applied to each family.

Figure 5 (main text) illustrates the impact of applying these updated frequency thresholds - derived from either the UK COVID-19 Genomics Study reference panel or gnomAD v4.1 (genomes) - on the number of variants prioritised relative to the standard pipeline, stratified by the assigned ancestry group of the probands these variants were identified in. An equivalent plot for candidate variants **outside** the applied virtual gene panels is provided in Figure A9.

**Stratified analyses by variant consequence and inheritance patterns**

We assessed whether the number and characteristics of variants prioritised during the UK100kGP rare disease programme - both those returned by the automated pipeline and those subsequently confirmed as diagnostic - varied across genetically inferred ancestry groups when stratified by molecular consequence and inferred inheritance pattern.

Variant consequences were grouped into the following categories based on Sequence Ontology terms recorded by the pipeline:

- **Protein-truncating variants**: stop-gained, frameshift, stop-lost, initiator codon, and start-lost (SO:0001587, SO:0001589, SO:0001578, SO:0001582, SO:0002012).
- **Canonical splice-site variants**: splice donor or acceptor sites (SO:0001574, SO:0001575)
- **Missense variants**: altered the amino acid sequence without introducing a stop (SO:0001583)
- **In-frame indels**: insertions or deletions that did not disrupt the reading frame (SO:0001821, SO:0001822)
- **Splice-region variants**: near but not within canonical splice sites (SO:0001630)
- **Varies by transcript**: reflecting transcript-specific differences in predicted consequence

Inheritance patterns were inferred from segregation tags generated by the pipeline and grouped as:

- **Dominant** *(including inherited autosomal dominant and imprinted)*
- **Recessive** (homozygous)
- **Recessive** (compound heterozygous)
- **De novo**
- **X-linked**
- **Mitochondrial**
- **Multiple plausible** *(more than one plausible model supported)*

These groupings were used to stratify two outcomes across genetically inferred ancestry groups: (1) the average number of prioritised variants per proband, and (2) the distribution of confirmed diagnostic variants (among those prioritised) across consequence types and inheritance patterns. Results are shown in Figures A4 and A5, respectively.

**Sensitivity analyses using updated virtual gene panel versions**

During the 100kGP analysis of primary clinical findings for rare diseases, virtual gene panels used to prioritise variants for manual review were selected by clinicians based on pre-test phenotypic information and applied using the then-current versions of Genomics England’s PanelApp resource (<https://panelapp.genomicsengland.co.uk/>), a dynamic, crowd-sourced platform that is continuously updated with evidence on gene–disease associations – including throughout the initial application of the automated variant prioritisation pipeline following sequencing.

To assess the robustness of ancestry-associated differences in pipeline performance - specifically the **sensitivity** and **positive predictive value (PPV)** of prioritised variants reported in the main text - we repeated key regression analyses using an updated set of virtual gene panels.

For this, we re-annotated variants previously returned outside of virtual gene panels using a set of updated PanelApp panel versions from March 1st, 2024 (<https://gitlab.com/genomicsengland/Data_Diversity_Public/rare-diseases-ancestry-paper>). We restricted to high-confidence (‘Green’) genes and matched panel names and inheritance models to those originally applied for each proband. This process identified **29 075** additional variants located within relevant disease genes, increasing the total number of prioritised variants to **85 717** across the cohort.

All multivariable regression models evaluating ancestry group differences in PPV and sensitivity (as described in the Statistical Analysis section of the main text) were re-run using this updated set of prioritised variants. Results are shown Figure A6.

**Table A1** – Numbers of probands assigned to each genetically inferred ancestry group, the reference populations merged to create them, the place of birth of individuals that comprised that reference group (taken from Prive et al. 2022), and comparisons with 16+1 and other reported parental ethnicities (>5 participants per ancestry group).

| **Ancestry group** | **#** | **Initial reference groups merged** | **Place of birth of reference individuals** | **Proband reported 16+1 ethnicities** | **Other reported parental ethnicities** |
| --- | --- | --- | --- | --- | --- |
| European | 23623 | Scandinavia, United Kingdom, Ireland, Finland, Europe South West, Europe South East, Europe North East, Italy | United Kingdom, Ireland, Denmark, Norway, Sweden, Germany, Netherlands, Iceland, Poland, Ukraine, Latvia, Germany, Kazakhstan, Lithuania, Romania, Serbia/Montenegro, Bosnia and Herzegovina, Bulgaria, Croatia, Hungary, Italy, Macedonia, Russia, Spain, Portugal, France, Gibraltar | British, Irish, Any other ethnic group, Any other mixed background, Any other White background, White and Asian, White and Black African, White and Black Caribbean | Albanian, American, Australian, British, Bulgarian, Canadian, Cyprus, Danish, Dutch, Eastern European, European, French, German, Greek, Greek Cypriot, Hungarian, Irish, Italian, Jewish, Kosovo, Latvian, Lithuanian, Maltese, New Zealand, Northern Irish, Norwegian, Polish, Portuguese, Romanian, Russian, Scottish, Slovakian, South African, Spanish, Swedish, Turkish, Ukraine, Welsh |
| South Asian | 3030 | Bangladesh, Pakistan, Sri Lanka | Bangladesh, Pakistan, Sri Lanka | Bangladeshi, British, Indian, Pakistani, White and Asian, Any other Asian background, Any other ethnic group, Any other mixed background | Afghanistan, Gujarati, Indian, Kashmir, Mauritius, Sri Lanka |
| Remaining participants | 941 | Unassigned, Japan, Philippines, South America | Japan, Philippines, Peru | African, Any other Asian background, Any other Black background, Any other ethnic group, Any other mixed background, Any other White background, British, Caribbean, White and Asian, White and Black African, White and Black Caribbean | Jamaican, Japanese, Philippines |
| West African | 679 | Africa West, Africa South | Zimbabwe, Uganda, South Africa, Zambia, Malawi, Tanzania, Congo, Nigeria, Ghana, Sierra Leone, Caribbean, Ivory Coast, Liberia, The Guianas, Togo | African, Any other Black background, Any other ethnic group, Caribbean, White and Black African, White and Black Caribbean | Barbados, Ghana, Jamaica, Jamaican, Nigeria |
| Middle Eastern | 452 | Middle East | Iraq, Iran, Turkey, Syria | Any other Asian background, Any other ethnic group, Any other mixed background, Any other White background, British, White and Asian | Cypriot, Cyprus, Iran, Iranian, Iraq, Iraqi, Kurdish, Lebanese, Romanian, Syrian, Turkish |
| Ashkenazi | 250 | Ashkenazi | United Kingdom, USA, Israel, Hungary, Ireland, France, Canada, Russia | Any other White background, British | Ashkenazi Jewish, German, Jewish, Russian |
| East African | 159 | Africa East | Somalia, Ethiopia, Sudan, Eritrea | African, Any other Black background, Any other ethnic group | Somalian, Sudan, Ethiopian, Eritrean |
| North African | 147 | Africa North | Algeria, Morocco, Egypt, Libya, Tunisia | Any other Asian background, Any other ethnic group, Any other White background | Algerian, Libya, Yemen |
| East Asian | 124 | Asia East | Hong Kong, Malaysia, China, Singapore, Vietnam, Indonesia, Taiwan, Thailand, Brunei, Macau (Macao) | Chinese, Any other Asian background, Any other ethnic group | Hong Kong |

**Table A2** – Distribution of standardised disease groups across the cohort by genetically inferred ancestry groups (see: <https://www.genomicsengland.co.uk/initiatives/100000-genomes-project/documentation> for disease data models).

|  | **European** | **East Asian** | **Middle Eastern** | **Ashkenazi** | **South Asian** | **North African** | **East African** | **West African** | **Remaining participants** |
| --- | --- | --- | --- | --- | --- | --- | --- | --- | --- |
| N total | 23623 | 124 | 452 | 250 | 3030 | 147 | 159 | 679 | 838 |
| **Disease group** |  |  |  |  |  |  |  |  |  |
| Neurology and neurodevelopmental disorders | 9729 (41.2%) | 44 (35.5%) | 162 (35.8%) | 99 (39.6%) | 1220 (40.3%) | 62 (42.2%) | 72 (45.3%) | 219 (32.3%) | 365 (43.6%) |
| Cardiovascular disorders | 2835 (12.0%) | 7 (5.6%) | 29 (6.4%) | 25 (10.0%) | 214 (7.1%) | 9 (6.1%) | <5 | 84 (12.4%) | 65 (7.8%) |
| Renal and urinary tract disorders | 2253 (9.5%) | 24 (19.4%) | 80 (17.7%) | 39 (15.6%) | 308 (10.2%) | 24 (16.3%) | 19 (11.9%) | 123 (18.1%) | 97 (11.6%) |
| Ophthalmological disorders | 1778 (7.5%) | 18 (14.5%) | 61 (13.5%) | 19 (7.6%) | 397 (13.1%) | 18 (12.2%) | 26 (16.4%) | 111 (16.3%) | 58 (6.9%) |
| Ultra-rare disorders | 1280 (5.4%) | 7 (5.6%) | 20 (4.4%) | 8 (3.2%) | 187 (6.2%) | 10 (6.8%) | 7 (4.4%) | 17 (2.5%) | 40 (4.8%) |
| Tumour syndromes | 1273 (5.4%) | <5 | 9 (2.0%) | 12 (4.8%) | 47 (1.6%) | <5 | <5 | 19 (2.8%) | 17 (2.0%) |
| Skeletal disorders | 599 (2.5%) | - | 12 (2.7%) | 8 (3.2%) | 66 (2.2%) | <5 | <5 | 10 (1.5%) | 21 (2.5%) |
| Endocrine disorders | 542 (2.3%) | 5 (4.0%) | 6 (1.3%) | 8 (3.2%) | 86 (2.8%) | <5 | <5 | 11 (1.6%) | 36 (4.3%) |
| Hearing and ear disorders | 460 (1.9%) | <5 | 18 (4.0%) | 5 (2.0%) | 103 (3.4%) | <5 | <5 | 11 (1.6%) | 29 (3.5%) |
| Haematological and immunological disorders | 510 (2.2%) | <5 | 9 (2.0%) | 5 (2.0%) | 44 (1.5%) | 5 (3.4%) | <5 | 12 (1.8%) | 10 (1.2%) |
| Metabolic disorders | 400 (1.7%) | <5 | 12 (2.7%) | <5 | 103 (3.4%) | <5 | <5 | 13 (1.9%) | 23 (2.7%) |
| Dysmorphic and congenital abnormality syndromes | 407 (1.7%) | <5 | 10 (2.2%) | <5 | 45 (1.5%) | <5 | <5 | 10 (1.5%) | 19 (2.3%) |
| Multiple disorders | 388 (1.6%) | <5 | 6 (1.3%) | <5 | 54 (1.8%) | - | 5 (3.1%) | 10 (1.5%) | 11 (1.3%) |
| Dermatological disorders | 208 (0.9%) | - | <5 | <5 | 67 (2.2%) | <5 | <5 | 16 (2.4%) | 18 (2.1%) |
| Respiratory disorders | 267 (1.1%) | - | <5 | 6 (2.4%) | 12 (0.4%) | - | <5 | <5 | 5 (0.6%) |
| Ciliopathies | 213 (0.9%) | <5 | 9 (2.0%) | <5 | 41 (1.4%) | <5 | <5 | <5 | <5 |
| Rheumatological disorders | 211 (0.9%) | <5 | <5 | <5 | 12 (0.4%) | <5 | <5 | <5 | 5 (0.6%) |
| Growth disorders | 137 (0.6%) | - | - | - | 10 (0.3%) | <5 | <5 | <5 | 6 (0.7%) |
| Gastroenterological disorders | 85 (0.4%) | - | <5 | <5 | 13 (0.4%) | <5 | - | <5 | 7 (0.8%) |
| Other disorders | 48 (0.2%) | <5 | - | - | <5 | <5 | <5 | <5 | <5 |

**Table A3**. Summary statistics of variant counts returned by the variant prioritisation pipeline genetically inferred ancestry group.

| **Outcomes** | **Overall** | **European** | **East Asian** | **Middle Eastern** | **Ashkenazi** | **South Asian** | **North African** | **East African** | **West African** | **Remaining participants** |
| --- | --- | --- | --- | --- | --- | --- | --- | --- | --- | --- |
| **Prioritised variants** |  |  |  |  |  |  |  |  |  |  |
| Total | 56642 | 40368 | 333 | 1063 | 323 | 8551 | 398 | 955 | 2159 | 2492 |
| Unique* | 37870 | 26320 | 327 | 1026 | 266 | 6858 | 387 | 858 | 1921 | 2294 |
| median (IQR) per proband | 1 (0–2) | 1 (0–2) | 1 (0–3) | 1 (0–3) | 0 (0–1) | 2 (0–4) | 2 (1–3) | 4 (1–8) | 2 (0–5) | 1 (0–4) |
| min-max per proband | 0–38 | 0–38 | 0–29 | 0–18 | 0–11 | 0–32 | 0–21 | 0–38 | 0–23 | 0–29 |
| >1 (n, %) | 17790 (60.5%) | 13665 (57.8%) | 90 (72.6%) | 326 (72.1%) | 124 (49.6%) | 2238 (73.9%) | 112 (76.2%) | 129 (81.1%) | 477 (70.3%) | 629 (66.8%) |
| **Candidate variants** (outside virtual panels) |  |  |  |  |  |  |  |  |  |  |
| Total | 5662419 | 4192244 | 33236 | 123066 | 39356 | 700966 | 36741 | 82551 | 241934 | 212325 |
| Unique* | 1955566 | 1379100 | 25324 | 86526 | 18893 | 348833 | 28082 | 46573 | 137909 | 147636 |
| median (IQR) per proband | 226 (43–283) | 224 (36–273) | 338 (66–386) | 364 (47–423) | 170 (116–211) | 209 (41–400) | 236 (45–416) | 620 (298–758) | 412 (208–480) | 235 (36–356) |
| min-max per proband | 0–1073 | 0–582 | 6–502 | 3–561 | 3–344 | 0–582 | 5–525 | 4–953 | 5–1073 | 1–980 |
| >1 (n, %) | 29398 (100.0%) | 23618 (100.0%) | 124 (100.0%) | 452 (100.0%) | 250 (100.0%) | 3028 (99.9%) | 147 (100.0%) | 159 (100.0%) | 679 (100.0%) | 941 (100.0%) |

**Table A4**. Summary statistics of outcomes recorded in the clinical scientist outcomes questionnaires for 29 405 probands stratified across genetically inferred ancestry groups.

| **Outcomes** | **European** | **East Asian** | **Middle Eastern** | **Ashkenazi** | **South Asian** | **North African** | **East African** | **West African** | **Remaining participants** |
| --- | --- | --- | --- | --- | --- | --- | --- | --- | --- |
| **Diagnostic Yield** |  |  |  |  |  |  |  |  |  |
| Diagnosis* recorded (%) | 4335 (18.4%) | 36 (29.0%) | 107 (23.7%) | 48 (19.2%) | 764 (25.2%) | 38 (25.9%) | 32 (20.1%) | 122 (18.0%) | 180 (19.1%) |
| No diagnosis recorded (%) | 19288 (81.6%) | 88 (71.0%) | 345 (76.3%) | 202 (80.8%) | 2266 (74.8%) | 109 (74.1%) | 127 (79.9%) | 557 (82.0%) | 761 (80.9%) |
| **VUS frequency** |  |  |  |  |  |  |  |  |  |
| VUS recorded (%) | 2232 (9.4%) | 13 (10.5%) | 37 (8.2%) | 8 (3.2%) | 375 (12.4%) | 8 (5.4%) | 22 (13.8%) | 74 (10.9%) | 120 (12.8%) |
| No VUS recorded (%) | 21391 (90.6%) | 111 (89.5%) | 415 (91.8%) | 242 (96.8%) | 2655 (87.6%) | 139 (94.6%) | 137 (86.2%) | 605 (89.1%) | 821 (87.2%) |

**Table A5.** Negative binomial mixed effects regression model output (prioritised variants as counts variable).

|  | **Number of prioritised variants returned** | | | |
| --- | --- | --- | --- | --- |
|  | **Unadjusted model** | | **Adjusted model** | |
|  | **IRR (95% CI)** | **p value** | **IRR (95% CI)** | **p value** |
| **Ancestry group** |  |  |  |  |
| European | 1 (ref) | — | 1 (ref) | — |
| Ashkenazi | 0.75 (0.62, 0.89) | 0.0015 | 0.75 (0.63, 0.89) | 0.001 |
| East African | 3.13 (2.60, 3.75) | <0.0001 | 2.77 (2.33, 3.29) | <0.0001 |
| East Asian | 1.52 (1.21, 1.90) | 0.0002 | 1.41 (1.14, 1.74) | 0.0015 |
| Middle Eastern | 1.34 (1.19, 1.51) | <0.0001 | 1.29 (1.15, 1.45) | <0.0001 |
| North African | 1.52 (1.24, 1.86) | <0.0001 | 1.38 (1.13, 1.68) | 0.0012 |
| Remaining participants | 1.47 (1.35, 1.60) | <0.0001 | 1.41 (1.30, 1.53) | <0.0001 |
| South Asian | 1.55 (1.47, 1.63) | <0.0001 | 1.47 (1.39, 1.56) | <0.0001 |
| West African | 1.88 (1.71, 2.07) | <0.0001 | 1.79 (1.63, 1.96) | <0.0001 |
| **Age at consent (per year)** |  |  |  |  |
| — | — | — | 0.99 (0.99, 0.99) | <0.0001 |
| **Sex** |  |  |  |  |
| Male | — | — | 1 (ref) | — |
| Female | — | — | 0.93 (0.90, 0.96) | <0.0001 |
| **Family structure** |  |  |  |  |
| Singleton | — | — | 1 (ref) | — |
| Family duo | — | — | 0.88 (0.84, 0.93) | <0.0001 |
| Family trio | — | — | 0.52 (0.49, 0.54) | <0.0001 |
| Other | — | — | 0.60 (0.56, 0.63) | <0.0001 |
| **Family disease affection** |  |  |  |  |
| Only proband affected | — | — | 1 (ref) | — |
| Unknown | — | — | 1.89 (1.79, 1.99) | <0.0001 |
| Family member affected | — | — | 0.94 (0.90, 0.98) | 0.0015 |
| **Predicted consanguinity** |  |  |  |  |
| Nonconsanguineous | — | — | 1 (ref) | — |
| Consanguineous | — | — | 1.25 (1.16, 1.34) | <0.0001 |
| Possibly consanguineous | — | — | 1.13 (0.98, 1.29) | 0.0865 |
| **Random effects** |  |  |  |  |
| • Disease group (σ) | 0.55 | — | 0.56 | — |
| • Handling GMC (σ) | 0.11 | — | 0.14 | — |

**Table A6**. Logistic mixed effects regression model output (diagnostic variant recorded = yes or no, as binary variable).

|  | **Diagnostic yield** | | | |
| --- | --- | --- | --- | --- |
|  | **Unadjusted model** | | **Adjusted model** | |
|  | **OR (95% CI)** | **p value** | **OR (95% CI)** | **p value** |
| **Ancestry group** |  |  |  |  |
| European | 1 (ref) | — | 1 (ref) | — |
| Ashkenazi | 1.16 (0.84, 1.61) | 0.3643 | 1.20 (0.87, 1.67) | 0.2641 |
| East African | 1.03 (0.69, 1.53) | 0.8767 | 0.92 (0.62, 1.38) | 0.6921 |
| East Asian | 1.76 (1.18, 2.62) | 0.0053 | 1.77 (1.18, 2.64) | 0.0054 |
| Middle Eastern | 1.28 (1.02, 1.60) | 0.0324 | 1.05 (0.83, 1.33) | 0.7085 |
| North African | 1.47 (1.01, 2.15) | 0.0452 | 1.18 (0.80, 1.74) | 0.4112 |
| Remaining participants | 1.02 (0.86, 1.20) | 0.8466 | 0.97 (0.82, 1.15) | 0.7366 |
| South Asian | 1.38 (1.26, 1.51) | <0.0001 | 1.10 (0.97, 1.23) | 0.1258 |
| West African | 0.92 (0.75, 1.13) | 0.4216 | 0.95 (0.77, 1.16) | 0.5925 |
| **Age at consent (per year)** |  |  |  |  |
| — | — | — | 1.00 (0.99, 1.00) | 0.0002 |
| **Sex** |  |  |  |  |
| Male | — | — | 1 (ref) | — |
| Female | — | — | 1.12 (1.05, 1.19) | 0.0003 |
| **Family structure** |  |  |  |  |
| Singleton | — | — | 1 (ref) | — |
| Family duo | — | — | 1.04 (0.93, 1.16) | 0.5325 |
| Family trio | — | — | 1.44 (1.29, 1.60) | <0.0001 |
| Other | — | — | 0.83 (0.74, 0.93) | 0.0016 |
| **Family disease affection** |  |  |  |  |
| Only proband affected | — | — | 1 (ref) | — |
| Unknown | — | — | 0.87 (0.76, 0.99) | 0.0376 |
| Family member affected | — | — | 1.72 (1.59, 1.87) | <0.0001 |
| **Predicted consanguinity** |  |  |  |  |
| Nonconsanguineous | — | — | 1 (ref) | — |
| Consanguineous | — | — | 1.48 (1.28, 1.71) | <0.0001 |
| Possibly consanguineous | — | — | 1.32 (1.01, 1.72) | 0.0404 |
| **Random effects** |  |  |  |  |
| • Disease group (σ) | 0.59 | — | 0.60 | — |
| • Handling GMC (σ) | 0.17 | — | 0.17 | — |

**Table A7.** Logistic mixed effects regression model output (variant level sensitivity; prioritised diagnostic variants / total diagnostic variants).

|  | **Sensitivity of prioritised variants** | | | |
| --- | --- | --- | --- | --- |
|  | **Unadjusted model** | | **Adjusted model** | |
|  | **OR (95% CI)** | **p value** | **OR (95% CI)** | **p value** |
| **Ancestry group** |  |  |  |  |
| European | 1 (ref) | — | 1 (ref) | — |
| Ashkenazi | 0.61 (0.31, 1.19) | 0.1478 | 0.55 (0.27, 1.09) | 0.0859 |
| East African | 1.44 (0.55, 3.76) | 0.4585 | 1.51 (0.57, 3.98) | 0.4076 |
| East Asian | 1.34 (0.58, 3.10) | 0.4974 | 1.35 (0.58, 3.15) | 0.4846 |
| Middle Eastern | 0.69 (0.44, 1.09) | 0.1118 | 0.76 (0.47, 1.23) | 0.2607 |
| North African | 0.94 (0.42, 2.08) | 0.8706 | 1.04 (0.46, 2.35) | 0.9264 |
| Remaining participants | 0.80 (0.56, 1.14) | 0.2142 | 0.85 (0.59, 1.21) | 0.3604 |
| South Asian | 1.01 (0.83, 1.22) | 0.9182 | 1.11 (0.86, 1.43) | 0.4065 |
| West African | 0.85 (0.54, 1.32) | 0.4589 | 0.85 (0.54, 1.33) | 0.4747 |
| **Age at consent (per year)** |  |  |  |  |
| — | — | — | 1.00 (1.00, 1.01) | 0.0447 |
| **Sex** |  |  |  |  |
| Male | — | — | 1 (ref) | — |
| Female | — | — | 1.01 (0.89, 1.15) | 0.8418 |
| **Family structure** |  |  |  |  |
| Singleton | — | — | 1 (ref) | — |
| Family duo | — | — | 0.78 (0.61, 1.00) | 0.0535 |
| Family trio | — | — | 0.68 (0.54, 0.86) | 0.0016 |
| Other | — | — | 0.48 (0.38, 0.62) | <0.0001 |
| **Family disease affection** |  |  |  |  |
| Only proband affected | — | — | 1 (ref) | — |
| Unknown | — | — | 1.28 (0.89, 1.84) | 0.1892 |
| Family member affected | — | — | 1.14 (0.96, 1.37) | 0.1451 |
| **Predicted consanguinity** |  |  |  |  |
| Nonconsanguineous | — | — | 1 (ref) | — |
| Consanguineous | — | — | 0.95 (0.70, 1.28) | 0.7274 |
| Possibly consanguineous | — | — | 1.15 (0.65, 2.03) | 0.6287 |
| **Random effects** |  |  |  |  |
| • Disease group (σ) | 0.41 | — | 0.50 | — |
| • Handling GMC (σ) | 0.58 | — | 0.56 | — |

**Table A8.** Logistic mixed effects regression model output (variant level PPV; prioritised diagnostic variants / total prioritised variants).

|  | **Positive predictive value (PPV) of prioritised variants** | | | |
| --- | --- | --- | --- | --- |
|  | **Unadjusted model** | | **Adjusted model** | |
|  | **OR (95% CI)** | **p value** | **OR (95% CI)** | **p value** |
| **Ancestry group** |  |  |  |  |
| European | 1 (ref) | — | 1 (ref) | — |
| Ashkenazi | 1.31 (0.93, 1.84) | 0.1222 | 1.45 (1.02, 2.06) | 0.0382 |
| East African | 0.29 (0.20, 0.42) | <0.0001 | 0.32 (0.22, 0.46) | <0.0001 |
| East Asian | 1.09 (0.76, 1.57) | 0.6272 | 1.16 (0.80, 1.68) | 0.4253 |
| Middle Eastern | 0.70 (0.56, 0.88) | 0.0021 | 0.68 (0.54, 0.86) | 0.0013 |
| North African | 0.74 (0.51, 1.07) | 0.1089 | 0.73 (0.50, 1.06) | 0.1017 |
| Remaining participants | 0.60 (0.50, 0.71) | <0.0001 | 0.59 (0.50, 0.70) | <0.0001 |
| South Asian | 0.71 (0.65, 0.77) | <0.0001 | 0.65 (0.58, 0.73) | <0.0001 |
| West African | 0.44 (0.36, 0.54) | <0.0001 | 0.47 (0.39, 0.57) | <0.0001 |
| **Age at consent (per year)** |  |  |  |  |
| — | — | — | 1.01 (1.00, 1.01) | <0.0001 |
| **Sex** |  |  |  |  |
| Male | — | — | 1 (ref) | — |
| Female | — | — | 1.18 (1.11, 1.25) | <0.0001 |
| **Family structure** |  |  |  |  |
| Singleton | — | — | 1 (ref) | — |
| Family duo | — | — | 1.25 (1.12, 1.39) | <0.0001 |
| Family trio | — | — | 2.69 (2.43, 2.97) | <0.0001 |
| Other | — | — | 1.46 (1.31, 1.63) | <0.0001 |
| **Family disease affection** |  |  |  |  |
| Only proband affected | — | — | 1 (ref) | — |
| Unknown | — | — | 0.48 (0.42, 0.55) | <0.0001 |
| Family member affected | — | — | 1.69 (1.57, 1.83) | <0.0001 |
| **Predicted consanguinity** |  |  |  |  |
| Nonconsanguineous | — | — | 1 (ref) | — |
| Consanguineous | — | — | 0.92 (0.80, 1.06) | 0.2499 |
| Possibly consanguineous | — | — | 0.91 (0.70, 1.17) | 0.4652 |
| **Random effects** |  |  |  |  |
| • Disease group (σ) | 1.01 | — | 1.01 | — |
| • Handling GMC (σ) | 0.15 | — | 0.17 | — |

**Table A9.** Logistic mixed effects regression model output (recorded VUS carrier status = yes or no, as binary variable).

|  | **Recorded VUS frequency (per proband)** | | | |
| --- | --- | --- | --- | --- |
|  | **Unadjusted model** | | **Adjusted model** | |
|  | **OR (95% CI)** | **p value** | **OR (95% CI)** | **p value** |
| **Ancestry group** |  |  |  |  |
| European | 1 (ref) | — | 1 (ref) | — |
| Ashkenazi | 0.63 (0.31, 1.31) | 0.2188 | 0.66 (0.32, 1.35) | 0.2530 |
| East African | 2.18 (1.34, 3.55) | 0.0016 | 1.90 (1.17, 3.10) | 0.0097 |
| East Asian | 1.31 (0.72, 2.42) | 0.3789 | 1.28 (0.69, 2.36) | 0.4308 |
| Middle Eastern | 1.23 (0.86, 1.75) | 0.2584 | 1.04 (0.72, 1.51) | 0.8342 |
| North African | 0.71 (0.34, 1.49) | 0.3685 | 0.62 (0.30, 1.29) | 0.2017 |
| Remaining participants | 1.49 (1.21, 1.84) | 0.0001 | 1.45 (1.17, 1.78) | 0.0005 |
| South Asian | 1.60 (1.41, 1.82) | <0.0001 | 1.35 (1.15, 1.59) | 0.0003 |
| West African | 1.48 (1.14, 1.92) | 0.0032 | 1.36 (1.05, 1.77) | 0.0217 |
| **Age at consent (per year)** |  |  |  |  |
| — | — | — | 1.00 (0.99, 1.00) | 0.0016 |
| **Sex** |  |  |  |  |
| Male | — | — | 1 (ref) | — |
| Female | — | — | 0.90 (0.83, 0.98) | 0.0167 |
| **Family structure** |  |  |  |  |
| Singleton | — | — | 1 (ref) | — |
| Family duo | — | — | 1.10 (0.95, 1.26) | 0.2065 |
| Family trio | — | — | 0.64 (0.55, 0.74) | <0.0001 |
| Other | — | — | 0.66 (0.56, 0.78) | <0.0001 |
| **Family disease affection** |  |  |  |  |
| Only proband affected | — | — | 1 (ref) | — |
| Unknown | — | — | 1.06 (0.90, 1.25) | 0.5035 |
| Family member affected | — | — | 1.10 (0.99, 1.23) | 0.0775 |
| **Predicted consanguinity** |  |  |  |  |
| Nonconsanguineous | — | — | 1 (ref) | — |
| Consanguineous | — | — | 1.44 (1.18, 1.77) | 0.0004 |
| Possibly consanguineous | — | — | 1.38 (0.96, 1.99) | 0.0846 |
| **Random effects** |  |  |  |  |
| • Disease group (σ) | 0.33 | — | 0.31 | — |
| • Handling GMC (σ) | 1.14 | — | 1.15 | — |

**Table A10**. Population allele frequency filters used by Genomics England’s variant prioritisation pipeline during the 100kGP (v5). Note, the South Asian population from gnomADv2 was excluded from these frequency filters due to a known technical error.

| **Database** | **Sequencing** | **Population** | **#** | **AF threshold Recessive** | **AF threshold** **Dominant** | **AF threshold** **Mitochondrial** |
| --- | --- | --- | --- | --- | --- | --- |
| gnomAD v2.1 | Exomes | African / African American (AFR) | 8,128 | 0.01 | 0.001 | _ |
| gnomAD v2.1 | Exomes | Admixed American (AMR) | 17,296 | 0.01 | 0.001 | _ |
| gnomAD v2.1 | Exomes | East Asian (EAS) | 9,197 | 0.01 | 0.001 | _ |
| gnomAD v2.1 | Exomes | Finnish European (FIN) | 10,824 | 0.01 | 0.001 | _ |
| gnomAD v2.1 | Exomes | Non-Finnish European (NFE) | 56,885 | 0.01 | 0.001 | _ |
| gnomAD v2.1 | Exomes | Ashkenazi (ASJ) | 5,040 | 0.01 | 0.001 | _ |
| gnomAD v2.1 | Exomes | Other (OTH) | 3,070 | 0.01 | 0.002 | _ |
| 1000 Genomes | Genomes | African (AFR) | 669 | 0.01 | 0.002 | 0.002 |
| 1000 Genomes | Genomes | Admixed American (AMR) | 352 | 0.01 | 0.002 | 0.002 |
| 1000 Genomes | Genomes | East Asian (EAS) | 515 | 0.01 | 0.002 | 0.002 |
| 1000 Genomes | Genomes | European (EUR) | 505 | 0.01 | 0.002 | 0.002 |
| 1000 Genomes | Genomes | South Asian (SAS) | 494 | 0.01 | 0.002 | 0.002 |
| Genomics England internal frequencies | Genomes | GEL | 6628 | 0.01 | 0.001 | _ |

**Table A11**. Population sizes and 95% filtering allele frequency (FAF) thresholds derived from the **UK COVID-19 Genomics Study** and retrospectively integrated into the automated variant prioritisation pipeline. Effective allele frequency (AF) thresholds shown here correspond to the observed AF at which the FAF falls below the filtering thresholds for dominant and recessive inheritance models. Thresholds were applied per ancestry group as shown.

| **Sequencing** | **Population** | **#** | **FAF threshold Dominant** | **Effective AF threshold Dominant** | **FAF threshold Recessive** | **Effective AF threshold Recessive** |
| --- | --- | --- | --- | --- | --- | --- |
| Genomes | European | 26923 | 0.001 | 0.001244 | 0.01 | 0.010753 |
| Genomes | South Asian | 2668 | 0.001 | 0.001874 | 0.01 | 0.012556 |
| Genomes | West African | 1185 | 0.001 | 0.002532 | 0.01 | 0.013781 |
| Genomes | Middle Eastern | 321 | 0.001 | 0.004673 | 0.01 | 0.018692 |
| Genomes | Ashkenazi | 276 | 0.001 | 0.005435 | 0.01 | 0.019685 |
| Genomes | East Asian | 264 | 0.001 | 0.005634 | 0.01 | 0.018939 |
| Genomes | East African | 105 | 0.001 | 0.009524 | 0.01 | 0.02551 |
| Genomes | North African | 72 | 0.001 | 0.013889 | 0.01 | 0.029851 |

**Table A12**. Population sizes and 95% filtering allele frequency (FAF) thresholds derived from the external **gnomADv4.1** reference database and retrospectively integrated into the automated variant prioritisation pipeline. Effective allele frequency (AF) thresholds shown here correspond to the observed AF at which the FAF falls below the filtering thresholds for dominant and recessive inheritance models. Thresholds were applied per ancestry group as shown.

| **Sequencing** | **Population** | **#** | **FAF threshold Dominant** | **Effective AF threshold Dominant** | **FAF_95_ threshold Recessive** | **Effective AF**  **threshold Recessive** |
| --- | --- | --- | --- | --- | --- | --- |
| Genomes & Exomes | European | 590035 | 0.001 | 0.001049 | 0.01 | 0.010152 |
| Genomes & Exomes | South Asian | 45548 | 0.001 | 0.001185 | 0.01 | 0.010565 |
| Genomes & Exomes | African/African American | 37545 | 0.001 | 0.001204 | 0.01 | 0.010631 |
| Genomes & Exomes | Admixed American | 30019 | 0.001 | 0.001228 | 0.01 | 0.010778 |
| Genomes & Exomes | East Asian | 22454 | 0.001 | 0.001266 | 0.01 | 0.010807 |
| Genomes & Exomes | Ashkenazi | 14804 | 0.001 | 0.001331 | 0.01 | 0.010998 |
| Genomes & Exomes | Middle Eastern | 3042 | 0.001 | 0.001808 | 0.01 | 0.012268 |

**Table A13** – Classified VUS recorded in the outcomes questionnaire in that were initially prioritised and returned to clinical scientists during the 100kGP return of primary rare disease findings; yet were filtered out using the ancestry group stratified FAF filters from the UK COVID-19 Study. Applied virtual gene panel name refers to the name of the PanelApp panel applied that this variant was originally prioritised under, whilst inheritance models refer to the underlying models (segregation patterns) that the variant was originally prioritised under. (GEL = Genomics England).

| **Variant** | **Consequence** | **Gene** | **Applied PanelApp virtual gene panel(s)** | **Inheritance model** | **UK COVID19 Study Maximum AF** | **gnomADv4**  **Maximum AF** | **GEL Pipeline Maximum AF** |
| --- | --- | --- | --- | --- | --- | --- | --- |
| chr1:11982112-AAAGT-A | Canonical splice-site (SO:0001575) | MFN2 | Hereditary neuropathy | Dominant | 0.033333 (East African) | _ | _ |
| chr1:155612177-T-C | Splice-region (SO:0001630) | MSTO1 | Congenital muscular dystrophy, Mitochondrial disorders | Recessive | 0.028986 (West African) | 0.017217 (African/African American) | 0.008325 (gnomADv2 AFR) |
| chr1:173909791-G-T | Missense (SO:0001583) | SERPINC1 | Inherited bleeding disorders | Dominant | 0.008061 (South Asian) | 0.007938 (South Asian) | 0.000679 (GEL) |
| chr1:201060785-C-T | Missense (SO:0001583) | CACNA1S | Congenital myopathy | Dominant | 0.007576 (East Asian) | 0.000334 (East Asian) | 0.000261 (gnomADv2 AFR) |
| chr2:178565038-T-C | Missense (SO:0001583) | TTN | Dilated Cardiomyopathy and conduction defects | Dominant | 0.014286 (East African) | 0.00024 (African/African American) | 0.000226 (GEL) |
| chr2:178636754-T-C | Missense (SO:0001583) | TTN | Dilated Cardiomyopathy and conduction defects | Dominant | 0.00625 (Middle Eastern) | 0.002489 (Middle Eastern) | 0.001 (1000G EUR) |
| chr2:190999619-C-A | Splice-region (SO:0001630) | STAT1 | Primary immunodeficiency | Dominant | 0.014286 (East African) | 0.000509 (African/African American) | 7.5e-05 (GEL) |
| chr2:203870802-G-A | Missense (SO:0001583) | CTLA4 | Primary immunodeficiency disorders | Dominant | 0.007788 (Middle Eastern) | 0.002309 (Middle Eastern) | 0.000379 (gnomADv2 NFE) |
| chr2:203870802-G-A | Missense (SO:0001583) | CTLA4 | Primary immunodeficiency | Dominant | 0.007788 (Middle Eastern) | 0.002309 (Middle Eastern) | 0.000379 (gnomADv2 NFE) |
| chr2:229808318-C-T | Missense (SO:0001583) | TRIP12 | Intellectual disability | Dominant | 0.019048 (East African) | 0.000134 (African/African American) | 7.5e-05 (GEL) |
| chr4:150872712-C-T | Missense (SO:0001583) | LRBA | A- or hypo-gammaglobulinaemia, Agranulocytosis, Combined B and T cell defect, Congenital neutropaenia, Multi-organ autoimmune diabetes, SCID | Recessive (compound het.) | 0.114286 (East African) | 0.002973 (Middle Eastern) | 0.0023 (1000G AFR) |
| chr6:152350676-A-G | Missense (SO:0001583) | SYNE1 | Limb girdle muscular dystrophy | Dominant | 0.019048 (East African) | 0.000574 (Ashkenazi) | 0.000813 (gnomADv2 ASJ) |
| chr7:156892978-C-T | Missense (SO:0001583) | LMBR1 | Skeletal dysplasia | Dominant | 0.001523 (European) | 0.001212 (European) | 0.000825 (gnomADv2 NFE) |
| chr8:10608115-G-C | Missense (SO:0001583) | RP1L1 | Posterior segment abnormalities | Dominant | 0.003858 (South Asian) | 0.003148 (South Asian) | 0.000679 (GEL) |
| chr8:10622700-C-T | Missense (SO:0001583) | RP1L1 | Retinal disorders | Dominant | 0.014286 (East African) | 0.000164 (Middle Eastern) | 3e-05 (gnomADv2 AMR) |
| chr9:127819664-C-T | Splice-region (SO:0001630) | ENG | Hereditary haemorrhagic telangiectasia | Dominant | 0.009524 (East African) | 0.000611 (Ashkenazi) | 0.000628 (gnomADv2 ASJ) |
| chr10:100993505-A-C | Missense (SO:0001583) | TWNK | Inherited white matter disorders, Mitochondrial disorders, Undiagnosed metabolic disorders | Dominant | 0.047619 (East African) | 0.001155 (Middle Eastern) | 8e-04 (1000G AFR) |
| chr11:61962823-G-A | Missense (SO:0001583) | BEST1 | Retinal disorders | Dominant | 0.014286 (East African) | 0.005774 (Middle Eastern) | 0.001094 (gnomADv2 OTH) |
| chr11:62692395-C-T | Missense (SO:0001583) | BSCL2 | Genetic epilepsy syndromes, Intellectual disability | Recessive | 0.038095 (East African) | 0.00076 (African/African American) | 0.0015 (1000G AFR) |
| chr13:114324819-A-G | Missense (SO:0001583) | CHAMP1 | Intellectual disability | Dominant | 0.02381 (East African) | 0.000493 (Middle Eastern) | NA (GEL) |
| chr15:31040113-C-T | Splice-region (SO:0001630) | TRPM1 | Retinal disorders | Recessive (compound het.) | 0.047619 (East African) | 0.002225 (African/African American) | 0.002222 (gnomADv2 AFR) |
| chr15:31061436-G-T | Splice-region (SO:0001630) | TRPM1 | Retinal disorders | Recessive (compound het.) | 0.047619 (East African) | 0.001852 (African/African American) | 0.002027 (gnomADv2 AFR) |
| chr16:2103764-G-A | Missense (SO:0001583) | PKD1 | Cystic kidney disease | Recessive (compound het.) | 0.012339 (European) | 0.011498 (European) | 0.008564 (gnomADv2 NFE) |
| chr16:2103764-G-A | Missense (SO:0001583) | PKD1 | Cystic kidney disease | Recessive (compound het.) | 0.012339 (European) | 0.011498 (European) | 0.008564 (gnomADv2 NFE) |
| chr16:2103764-G-A | Missense (SO:0001583) | PKD1 | Cystic kidney disease, Rare multisystem ciliopathy disorders | Recessive (compound het.) | 0.012339 (European) | 0.011498 (European) | 0.008564 (gnomADv2 NFE) |
| chr16:2103764-G-A | Missense (SO:0001583) | PKD1 | Cystic kidney disease | Recessive (compound het.) | 0.012339 (European) | 0.011498 (European) | 0.008564 (gnomADv2 NFE) |
| chr16:71067297-T-C | Missense (SO:0001583) | HYDIN | Primary ciliary disorders | Recessive (compound het.) | 0.066327 (East African) | 0.006721 (African/African American) | 0.004074 (GEL) |
| chr16:89283717-C-T | Missense (SO:0001583) | ANKRD11 | Intellectual disability | Dominant | 0.014286 (East African) | 0.000347 (Middle Eastern) | _ |
| chr17:38328267-C-T | Missense (SO:0001583) | GPR179 | Posterior segment abnormalities | Recessive (compound het.) | 0.033333 (East African) | 0.00033 (Middle Eastern) | _ |
| chr17:38328600-C-T | Missense (SO:0001583) | GPR179 | Posterior segment abnormalities | Recessive (compound het.) | 0.033333 (East African) | 0.00033 (Middle Eastern) | _ |
| chr20:62836323-C-T | Missense (SO:0001583) | COL9A3 | Multiple Epiphyseal Dysplasia, Unexplained skeletal dysplasia | Dominant | 0.009524 (East African) | 0.00099 (Middle Eastern) | 0.001 (1000G EUR) |
| chrX:101397946-T-C | Missense (SO:0001583) | GLA | Hypertrophic Cardiomyopathy, Undiagnosed metabolic disorders | Dominant | 0.005048 (South Asian) | 0.004608 (South Asian) | 0.000528 (GEL) |
| chrX:154031004-A-G | Missense (SO:0001583) | MECP2 | Intellectual disability | Dominant | 0.036232 (East African) | 0.000157 (African/African American) | 0.000247 (gnomADv2 OTH) |

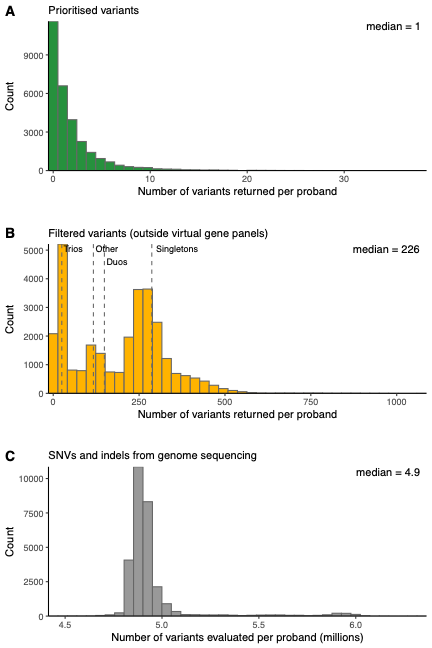

**Figure A1.** Histograms showing the numbers of variants returned per probands per variant type (A) prioritised variants. (B) Candidate variants returned outside of the virtual gene panels (C) Total SNVs and indels evaluated by the pipeline.

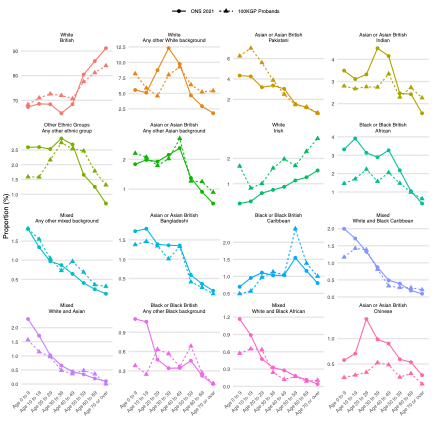

**Figure A2.** Proportion of probands in the UK100kGP reporting to each of 16+1 ethnic group categories (excluding individuals whose ethnic group was ‘Not known’ or was not stated) (dashed line) alongside the proportion of individuals self-reporting to each ethnic group as recorded in the ONS 2021 Census for England, stratified by age (in decades).

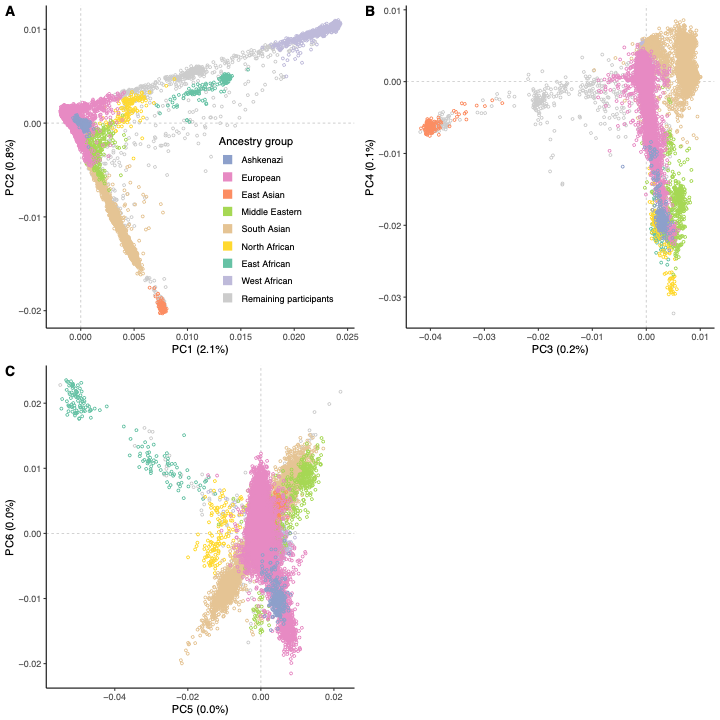

**Figure A3.** Principal component analysis (PCA) of genotypes, showing the top six genotype-derived principal components (PCs), coloured by genetically inferred ancestry group. (A) PC1 vs PC2, (B) PC3 vs PC4, and (C) PC5 vs PC6. The percentage of total genetic variance explained by each PC is indicated on the corresponding axis.

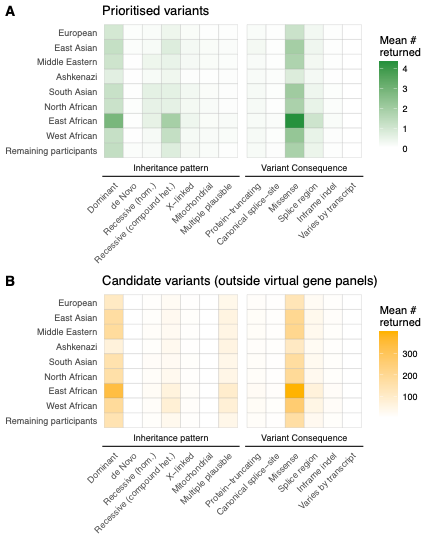

**Figure A4.** Heatmaps showing the mean number of variants returned by the automated variant prioritisation pipeline per proband by genetically inferred ancestry group and by inferred inheritance pattern and molecular consequence: (A) variants prioritised within the applied virtual gene panels, and (B) candidate variants outside those panels.

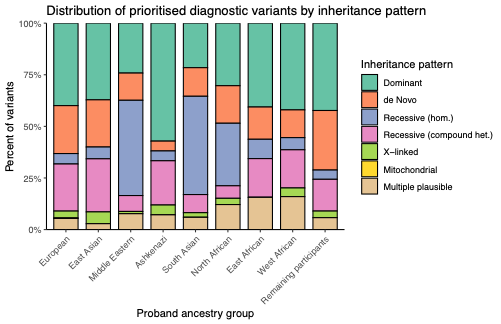

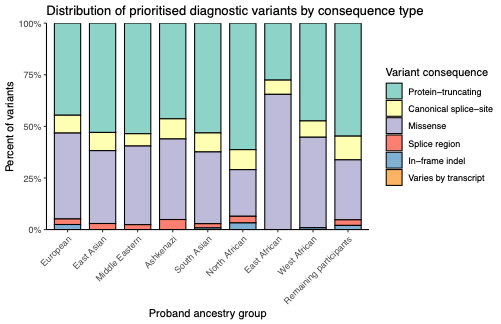

**Figure A5.** Stacked bar chart showing, for each genetically inferred ancestry group, the proportion of reported diagnostic variants prioritised by the pipeline that fall into (**A**) each inferred inheritance pattern and (**B**) each variant consequence category.

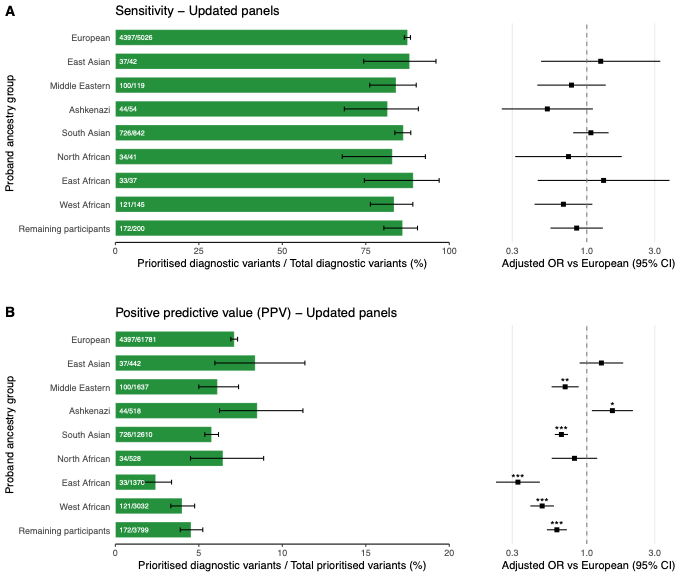

**Figure A6. Sensitivity and positive predictive value of prioritised variants (using updated virtual gene panels), stratified by genetically inferred ancestry group.** **(A)** Left: proportion of reported diagnostic SNVs and indels intersecting with variants retrospectively prioritised using the updated virtual gene panels (sensitivity) shown as green bars with numerator/denominator labels inside each bar and 95% Clopper–Pearson CIs (whiskers). Right: adjusted‐OR forest plot. **(B)** Left: proportion of these updated panel SNVs and indels reported as diagnostic (PPV) shown as green bars with numerator/denominator labels inside each bar and 95% Clopper–Pearson CIs (whiskers). Right: adjusted-OR forest plot. Estimates of adjusted OR vs European are from multivariable mixed‐effects logistic regression models adjusted for age, sex, family structure, family‐member affection status, and predicted consanguinity, with random intercepts for disease category and the handling GMC. *** p<0·001; ** p<0·01; * p<0·05).

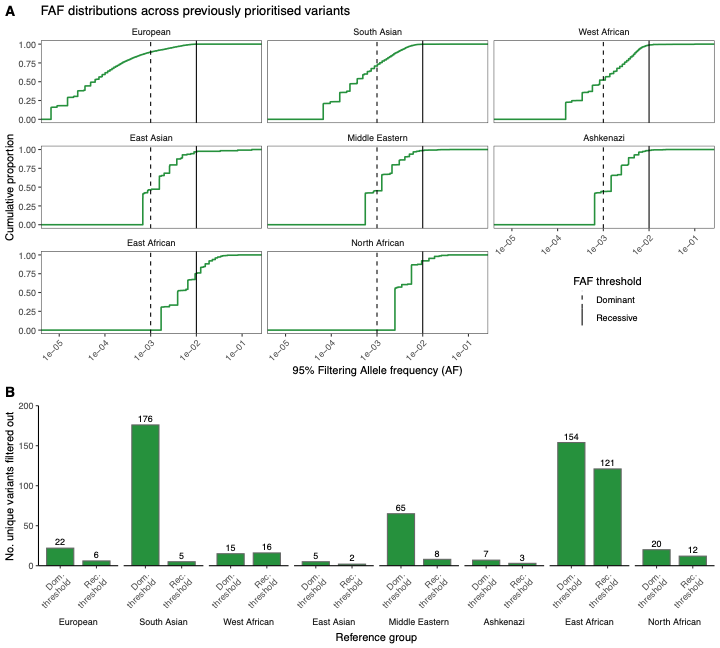

**Figure A7.** (A) Ancestry group specific FAF distributions from the UK COVID-19 Genomics Study reference panel for all previously prioritised variants in the 100 000 Genomes Project rare diseases cohort. FAF thresholds are shown for dominant and recessive inheritance models annotated by the pipeline (B) For each reference ancestry group, the number of (unique) prioritised variants that were filtered out in each of the reference ancestry groups at each threshold.

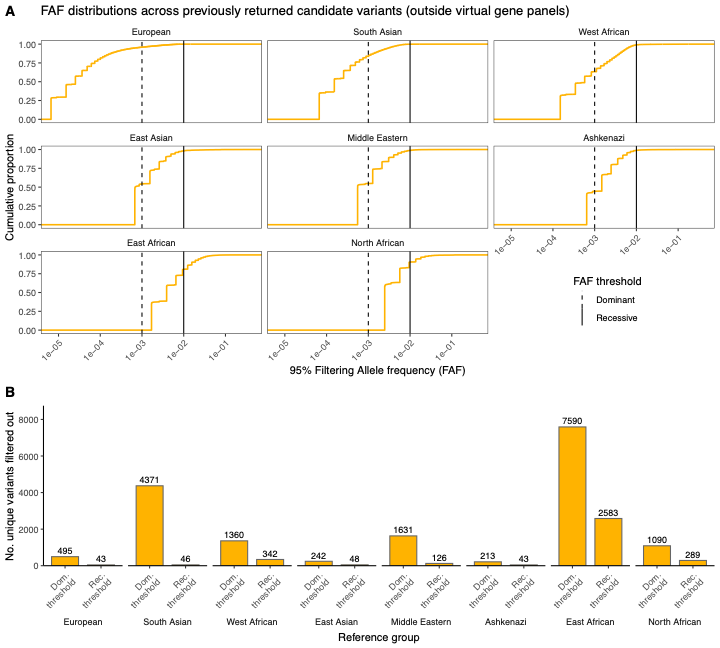

**Figure A8.** (A) Ancestry group specific FAF distributions from the UK COVID-19 Genomics Study reference panel for all previous candidate variants outside the virtual panels in the 100 000 Genomes Project rare diseases cohort. FAF thresholds are shown for dominant and recessive inheritance models annotated by the pipeline (B) For each reference ancestry group, the number of (unique) candidate variants outside panels that were filtered out in each of the reference ancestry groups at each threshold.

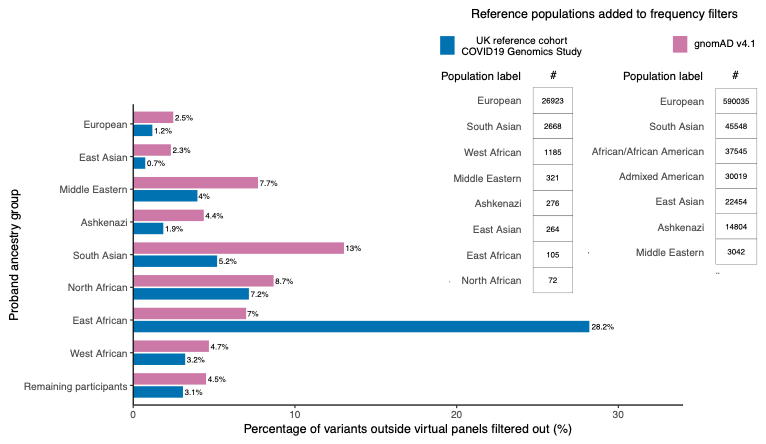

**Figure A9.** **Impact of additional population frequency filters on candidate variants outside virtual panels across eight ancestry groups.** Blue bars show the percentage of candidate variants outside the applied panels removed for each ancestry group, when additional allele frequency thresholds estimated from the UK COVID-19 Genomics Study (blue) were added to standard pipeline allele frequency filters. Magenta bars show the same for allele frequencies derived from seven reference groups in gnomADv4.1 (magenta). Sample sizes for each labelled reference group are shown in the inset tables.
